# Vector2Variant: Discovery of Genetic Associations from ML Derived Representations without Phenotype Engineering

**DOI:** 10.64898/2026.04.10.26350624

**Authors:** Matt Sooknah, Ramprakash Srinivasan, Sivaramakrishnan Sankarapandian, Zhenghao Chen, Jun Xu

## Abstract

Genome-wide association studies (GWAS) have transformed our understanding of human biology, but are constrained by the need for predefined phenotypes. We introduce Vector2Variant (V2V), a general-purpose framework that transforms any set of high-dimensional measurements (such as machine learning embeddings) into a genome-wide scan for associations, without requiring rigid specification of a phenotype. Rather than testing genetic variants against single traits, V2V finds the axis in multivariate space along which carriers and non-carriers maximally differ, and produces a continuous “projection phenotype” that can be interpreted by association with disease labels. The projection phenotypes correlate with orthogonal clinical biomarkers never seen during training, suggesting the learned axes capture biologically meaningful variation. We applied V2V to imaging, timeseries, and omics modalities in the UK Biobank and recovered established biology (like the role of CASP9 in renal failure) without the need for targeted measurements, alongside novel associations including a frameshift variant in LRRIQ1 (potentially protective for cardiovascular disease). V2V is computationally efficient at genome-wide scale, producing summary statistics and disease associations that facilitate target prioritization without the need for phenotype engineering.

## Main

Genome-wide association studies (GWAS) are an essential tool for systematically linking genetic variants to observable traits [1]. However, genetic analyses have traditionally been constrained by our ability to define and quantify informative phenotypes. While genomic data offers a comprehensive and hypothesis-free survey of natural variation, phenotypic data from dense modalities is typically reduced to a library of handcrafted features, such as organ volumes in magnetic resonance imaging (MRI) or specific waveforms like P/QRS complexes in electrocardiograms (ECG) [2–5]. These features are typically defined through careful application of domain knowledge (a process we refer to as *phenotype engineering*). These reductionist approaches work well when data modalities are well understood and there is sufficient prior knowledge of informative features that can be extracted from the data. The disadvantages of pre-defined features are that (a) they may require expensive expert annotation or development of specialized annotation pipelines to scale to large sample sizes, (b) they discard a large fraction of the information contained in high-dimensional biological measurements that is not well captured by human defined features, and (c) they preclude the use of emerging data modalities where we have weaker prior knowledge on informative ways to featurize the data. These limitations have become increasingly restrictive, particularly as biobanks scale to include multiple dense, high-dimensional data modalities ranging from static 3D whole-body imaging and dynamic cardiac videos to time-series electrocardiograms and plasma proteomics [6].

In our work, we present an approach which first identifies systematic effects of genetic variation as they manifest in high-dimensional representations of biological states and then link these effects to associations with disease risk or known disease biomarkers. Our work is motivated by recent advances in self-supervised machine learning (ML) which have enabled the automated extraction of rich, high-dimensional embeddings (also called *representations*) that capture the intrinsic structure of dense data modalities [7, 8]. The primary advantage of these ML representations is that they are entirely defined by the data, and can potentially capture subtle signals that are not apparent to human perception [9, 10]. However, the best way to utilize these embeddings for genetic discovery remains an open problem. Standard approaches typically rely on dimensionality reduction or clustering [11, 12], which are not guaranteed to capture the variety of unique biological processes, or use embeddings to predict existing phenotypes, which suffers from the same limitations of manual phenotype definition.

Our work draws inspiration from prior approaches which identify genetic factors that are associated with known biomarkers of disease or function (e.g., estimated glomerular filtration rate), thereby linking these with associated diseases (e.g., renal failure) [13]. However, the key idea behind our approach is to reverse the inference direction: rather than searching for genetic association with a fixed set of phenotypes, we infer the effects of a genetic variant by searching for directions in the multivariate phenotypic representation space that optimally separate individuals based on their genotype. This approach relies on the observation that if a genetic variant exerts a consistent biological effect that is reflected in a data modality of interest, it should induce a corresponding systematic shift in a sufficiently expressive representation of the data. We can therefore test for genetic variants that have consistent effects by asking whether genotype classes are separable in our representation space: if they are, the direction of maximal separation defines a *de novo* phenotype capturing the variant’s expected effects as it manifests in our data modality; if they are not, we conclude the variant has no detectable effect within that modality.

Here, we introduce Vector2Variant (V2V), a scalable framework for identifying genetic associations directly from multivariate representations of dense biological data (Fig. 1). For each genetic variant, our framework identifies the specific combination of features that separate carriers from non-carriers, producing a continuous “projection score” for every individual in the study that quantifies how closely their multivariate data aligns with the expected effect of the variant. To understand the biological meaning of these scores, we use a framework similar to a phenome-wide association study (PheWAS) [14]. While a traditional PheWAS tests the association between genotype dosage and a list of traits, we replace the dosage with the continuous projection score derived from each variant. This approach, which we term a Projection-Phenome Association Study (ProjWAS), reveals which diseases are most strongly associated with the organ-specific features that separate carriers from non-carriers of a particular allele.

**Fig. 1.**
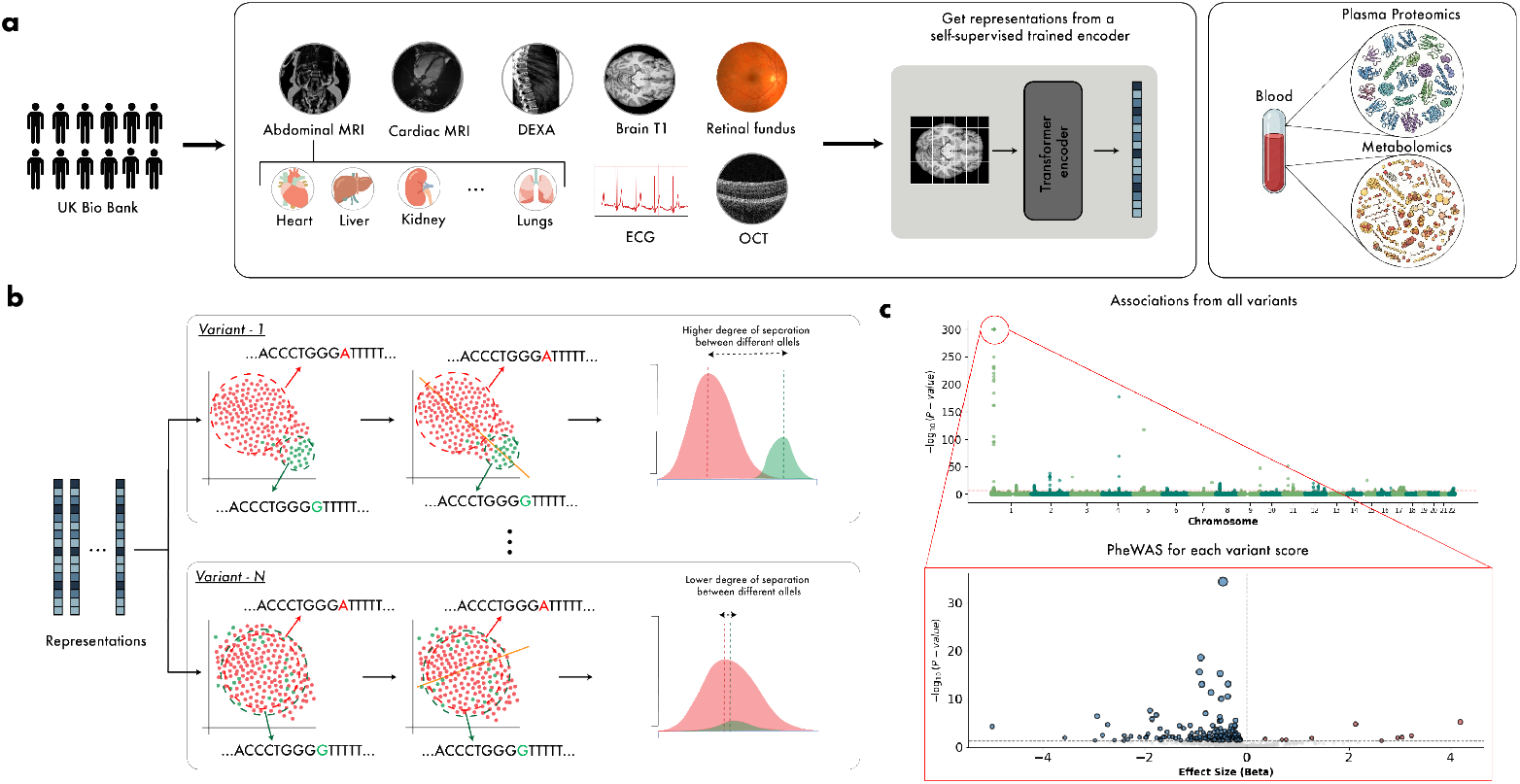
Vector2Variant framework for phenotype-free genetic discovery. **a**, High-dimensional data across modalities (MRI, video, ECG, omics) are converted into compact embedding vectors using self-supervised models such as masked autoencoders. **b**, For each genetic variant, V2V trains a linear classifier to separate carriers (red) from non-carriers (green) in embedding space. When genotype classes are separable, projection onto the discriminant axis produces distinct score distributions (right, top); when inseparable, distributions overlap (right, bottom). **c**, Genome-wide association testing across all variants produces a Manhattan plot of projection phenotype associations. For significant loci, the projection scores are carried forward into a projection-phenome association study (ProjWAS) against clinical diagnoses to interpret the biological meaning of each learned axis. All panels show schematic illustrations.

V2V has several advantages: (a) it is sensitive to effects that are not well captured by existing features or apparent to human perception and intuition, (b) it enables use of data modalities where we don’t have good existing feature extraction pipelines, (c) since our phenotypic score is defined directly by the data, we bypass the immense multiple hypothesis testing burden typically associated with high-dimensional data, and (d) by scoring the entire cohort on how much they “resemble” variant carriers, we borrow statistical power from individuals who phenocopy a true carrier. This can provide greater power to detect subtle signals, particularly for rare variants. Our method complements existing workflows for genetic discovery, potentially serving as a “first pass” to identify promising modalities and signals in a cohort, which could lead to development of more targeted phenotypes for GWAS follow-up.

To illustrate the utility of our method, we applied our framework to a diverse array of data modalities from the UK Biobank (UKBB) [6] spanning abdominal MRI, brain MRI, retinal imaging (OCT, retinal fundus), cardiac video MRI, and 12-lead ECG. In total, we analyzed embeddings derived from 17 distinct modalities across 10 organ systems (*n*_*mean*_ = 77, 223 participants per modality), identifying a total of 5, 304 significant locus-modality associations, of which 474 are novel (relative to the GWAS Catalog [15]). Additionally, we extended the method to high-dimensional omics data, analyzing plasma proteomics [16] (PPP, *n* = 48, 581) and metabolomics [17] (NMR, *n* = 248, 183). For omics data, we apply our method directly to the measured features (251 metabolites in NMR, 2913 proteins in PPP) without the need for an embedding step.

Our method recovers from raw data alone many known associations that previously required collection of targeted functional phenotypes or additional feature extraction (Fig. 2). For example, the *CASP9*-renal failure association [13] emerges from kidney MRI without eGFR measurement, *ADGRG6*-COPD [18] from lung MRI instead of spirometry measurements, *IL11*-hyperostosis [19] from spine DXA without pathology scoring, and *SCN10A*-conduction abnormalities [20] from raw ECG waveforms without QRS interval extraction.

**Fig. 2.**
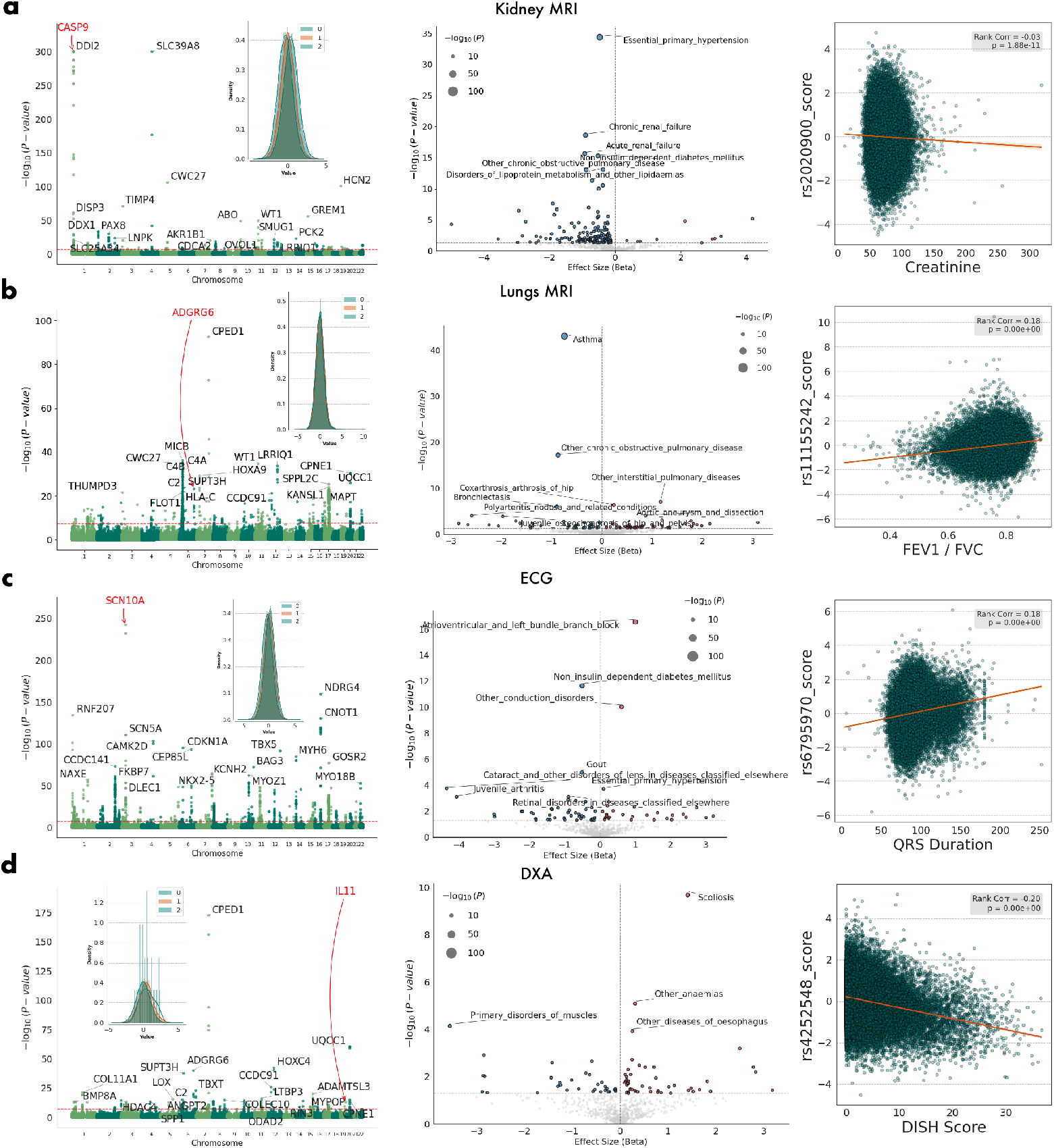
Projection phenotypes recover established biological associations without manual feature engineering. **Left and middle columns**, Manhattan plots (left) and corresponding Pro- jWAS volcano plots (middle) for the variant highlighted in the Manhattan plots detailing the genetic and clinical associations for four representative variant-specific projection phenotypes. **a**, Kidney MRI embeddings highlight the *CASP9* locus; the ProjWAS signal, driven by latent tissue features, strongly associates with hypertension and chronic renal failure, replicating eGFR-based findings. **b**, Lung MRI embeddings highlight the *ADGRG6* variant. The projection phenotype successfully captures morphological signatures of airflow limitation and COPD despite relying solely on fat and water channels from lung MRI. **c**, ECG embeddings highlight an *SCN10A* variant associated with cardiac conduction abnormalities. **d**, Lateral spine DXA embeddings highlight an *IL11* variant strongly associated with DISH. **Right column**, Scatter plots validating the derived projection scores against established orthogonal measurements—serum creatinine, liver iron concentration, ECG QRS duration, and DISH score—displaying corresponding correlation coefficients and *P* -values.

V2V learns distinct projection phenotypes for the same variant in different modalities, revealing organ-specific disease signatures in genes like *SH2B3* [21] and *SLC39A8* [22]. In downstream ProjWAS, the projection phenotypes for each organ system preferentially associate with biologically concordant disease categories (e.g., brain MRI with nervous system disorders, kidney MRI with genitourinary conditions), while systemic conditions like metabolic disorders are captured across almost all organs.

Beyond recovering known biology, V2V reveals novel genetic associations. For instance, a frameshift variant in *LRRIQ1*, a gene with established roles in fertility and reproductive biology [23, 24], exhibits a consistent protective cardiovascular signature in ProjWAS across kidney, liver, heart, and lung embeddings, with top disease associations spanning cystic kidney disease, hypertension, atrial fibrillation, and ischemic heart disease. The convergence of this signal across four independent organ systems suggests a broader role in vascular health than previously recognized. Together, these results demonstrate that V2V provides a unified strategy to discover genetic associations from high-dimensional data and generate interpretable clinical hypotheses to aid in target discovery.

## Results

### The Vector2Variant Framework

The V2V framework operates on a matrix of input features *𝒳* ∈ ℝ^*N ×D*^, where each individual is represented by a *D*-dimensional vector derived either from self-supervised models applied to raw data (images, videos, time series) or directly from molecular assay measurements (proteomics, metabolomics). The framework proceeds in three stages: learning a variant-specific projection axis, evaluating its statistical significance on held-out data, and interpreting the resulting phenotype through clinical association testing (ProjWAS, Fig. 1).

For each genetic variant, we seek a specific axis in the multivariate space that best captures the biological perturbation driven by that variant. We train a linear classifier (Linear Discriminant Analysis [25]) to distinguish carriers of the alternate allele from homozygous reference individuals. This identifies a vector **ŵ**_*v*_, the normal to the decision boundary, that defines the axis of maximal separation between genotype classes, onto which individuals are projected to produce a continuous “projection phenotype”. We employ two training strategies: a “Homozygous Contrast” model which trains on the genotypic extremes (homozygous reference vs. homozygous alternate), and a “Dominant Contrast” model which trains on reference vs. carriers (heterozygous + homozygous alternate). Unless otherwise specified, results presented utilize the Dominant Contrast model.

To prevent overfitting, a critical risk when the number of embedding dimensions approaches the number of variant carriers, we employ a strict cross-validation scheme. The projection axis ŵ_*v*_ is learned solely on a training subset. We then project a held-out validation set containing all three genotype classes (reference, heterozygous, and alternate) onto this axis to generate continuous scores (y = 𝒳_val_ŵ_*v*_). This scalar score quantifies the degree to which an individual exhibits the biological perturbation associated with that variant in the specified high-dimensional feature space. Statistical significance is assessed by testing whether these validation scores scale linearly with allele count *g* ∈ {0, 1, 2} via linear regression, ensuring the learned axis captures a genuine phenotypic gradient rather than overfitting to the training partition. We aggregate results across cross-validation folds using the harmonic mean of the resulting p-values. A summary of primary associations across 17 organ modalities is reported in Table 1.

**Table 1.** Number of significant loci discovered in each modality using different contrasts, along with totals for different modality groupings. Asterisk denotes a primary modality, selected as the “representative” for that organ system in select analyses (e.g. ProjWAS category summarization). See Methods for detailed counting methodology.

| Modality (* = primary) | System | Homozygous | Dominant | Combined |
| --- | --- | --- | --- | --- |
| Spine DXA* | Bone | 61 | 100 | 122 |
| Brain (T1)* | Brain | 675 | 921 | 1222 |
| Heart Video (short axis)* | Cardiac | 630 | 795 | 989 |
| Heart Video (long axis) | Cardiac | 302 | 386 | 509 |
| Heart Image (Dixon scan) | Cardiac | 251 | 326 | 428 |
| ECG | Cardiac | 184 | 265 | 310 |
| Aorta | Cardiac | 104 | 118 | 178 |
| OCT (right eye) | Eye | 88 | 254 | 281 |
| OCT (left eye)* | Eye | 98 | 260 | 287 |
| Retinal Fundus | Eye | 98 | 137 | 157 |
| Right Kidney* | Kidney | 73 | 107 | 135 |
| Left Kidney | Kidney | 62 | 100 | 118 |
| Liver* | Liver | 90 | 108 | 147 |
| Lungs* | Lungs | 124 | 182 | 242 |
| Pancreas* | Pancreas | 22 | 36 | 48 |
| Spleen* | Spleen | 47 | 71 | 89 |
| Bladder | Bladder | 15 | 30 | 42 |
| **Total (all organs)** | - | **2924** | **4196** | **5304** |
| **Total (primary organs)** | - | **1820** | **2580** | **3281** |

Finally, to interpret the biological meaning of a significant projection y, we test its association with ICD10 disease labels *d*_1_ … *d*_*N*_ ∈ 0, 1, henceforth referred to as a Projection-Phenome Association Study (ProjWAS). This reveals which diseases are significantly associated with the organ-specific features that are most informative for classifying carriers vs non-carriers of the variant. Since increasing projection score is analogous to increasing allelic dosage, the effect size of the disease association enables separation of protective vs deleterious effects. While we focus on disease associations, the method generalizes to any set of phenotypic traits of interest (biomarkers, lifestyle factors), which could be used to further interpret the variant score. The implementation scales to millions of variants across hundreds of features and many dense modalities by leveraging GPU acceleration.

### V2V recovers known genetic associations without phenotype engineering

To demonstrate the efficacy of our embedding-based approach, we highlight four established genetic associations recovered by our method without the use of explicitly derived quantitative phenotypes (for novel associations, see Section 1). Previously, these quantitative traits (e.g., FEV1/FVC) have often required measurement via different modalities or complex phenotypic derivations.

The estimated glomerular filtration rate (eGFR) is a clinical index that quantifies the volume of blood filtered per minute by the renal glomerular capillaries, serving as the primary metric for staging chronic kidney disease [26]. Genome-wide association studies (GWAS) of large European-ancestry cohorts have linked the *CASP9* locus to eGFR, and previous studies [13] have implicated *CASP9* as a causal gene for kidney dysfunction through the integration of GWAS and human kidney expression data.

While eGFR is conventionally derived from serum creatinine measurements, applying our method to segmented kidney embeddings from abdominal MRI in the UK Biobank (UKB) (Fig. 2) identifies *CASP9* as the most significant locus (lead variant rs2020900, *P <* 1 *×* 10^*−*300^). Subsequent ProjWAS of the variant score identifies *chronic renal failure* (*β* = *−* 0.90, *P* = 2.29 *×* 10^*−*19^) among the top associations. Notably, this association is recovered entirely from imaging features without an explicitly defined phenotype. To our knowledge, the association between *CASP9* and chronic renal failure has not been previously established using imaging as a primary readout. Furthermore, kidney embeddings from homozygous reference and alternative allele carriers exhibit a high degree of linear separability, indicating that the embeddings reflect phenotypic changes associated with these variants.

Chronic obstructive pulmonary disease (COPD) is a polygenic condition historically defined by spirometric airflow limitation, particularly the FEV1/FVC ratio. While previous GWAS using these standard metrics have linked the *ADGRG6* locus to pulmonary function [18], our approach maps this genetic architecture using an atypical imaging modality, specifically MRIs instead of CT scans. As shown in Fig. 2, we identify *ADGRG6* as a top signal (rs11155242, *P* = 5.11 *×* 10^*−*15^) utilizing latent embeddings extracted exclusively from lung MRIs. ProjWAS of the rs11155242 variant score highlights significant inverse associations with *asthma* (*β* = *−*0.74, *P* = 8.95 *×*10^*−*44^) and *other chronic obstructive pulmonary disease* (*β* = *−*0.87, *P* = 6.58 *×* 10^*−*18^). Corroborating this link, the imaging-derived variant score exhibits a positive correlation with spirometric FEV1/FVC (*ρ* = 0.18, *P <* 1 × 10^*−*300^) supporting the association with COPD. This demonstrates that MRI embeddings can capture morphological signatures related to COPD and recover genetic associations in the absence of explicit feature training.

The QRS duration on an electrocardiogram (ECG) is a fundamental quantitative trait reflecting ventricular depolarization time, prolonged intervals serve as potent clinical biomarkers for structural heart disease, arrhythmias, and sudden cardiac death [27]. Multiple studies [3, 28] have linked QRS prolongation to genetic variants at the *SCN10A* locus. Applying our framework to embeddings derived directly from raw UKB ECG waveforms (see Methods) identifies *SCN10A* as a top locus (lead variant rs6795970, *P* = 4.48 *×* 10^*−*243^). Importantly, this association is detected without explicitly deriving QRS duration, relying instead on the implicit information captured by the embeddings. ProjWAS of the rs6795970 variant score yields *atrioventricular and left bundle branch block* as the top association (*β* = 1.00, *P* = 2.89 *×* 10^*−*17^), which is consistent with existing literature.

Previous genomic insights into the role of *IL11* in bone homeostasis have largely depended on the construction of complex, ML derived phenotypes. For instance, [19] utilized a sophisticated machine learning pipeline to calculate a continuous Diffuse Idiopathic Skeletal Hyperostosis (DISH) score, revealing an association between the protein-coding variant rs4252548 and spinal hyperostosis using lateral dual-energy X-ray absorptiometry (DXA) scans in the UKB. Applying our method to embeddings derived from this same set of lateral DXA scans demonstrates that reference and alternate alleles for rs4252548 at the *IL11* locus are linearly separable with high statistical significance (*P* = 4.75*×*10^*−*14^). Consequently, our self-supervised embedding approach successfully recovers the association between *IL11* and bone traits, circumventing the need for specialized machine learning pipelines to derive quantitative phenotypes.

### Projection scores correlate with established clinical biomarkers and capture expected morphology

Our proposed method generates two primary outputs for each genetic variant: a projection phenotype and its corresponding LDA weights. To determine whether these projection phenotypes capture the underlying biological features of interest, we evaluated their correlation with the established orthogonal clinical measurements that originally defined these gene-disease associations. As illustrated in Fig. 2 (right column), the variant-specific projection scores exhibit significant correlations with relevant clinical biomarkers. Specifically, the rs2020900 projection score at the *CASP9* locus correlates with serum creatinine (*ρ* = *−*0.03, *P* = 1.88 *×* 10^*−*11^), the rs11155242 score at the *ADGRG6* locus with FEV1 / FVC (*ρ* = 0.18, *P <* 1 *×* 10^*−*300^), the rs4252548 score at the *IL11* locus with the DISH score (*ρ* = *−*0.20, *P <* 1 *×* 10^*−*300^), and the rs6795970 score at the *SCN10A* locus with ECG QRS duration (*ρ* = 0.18, *P <* 1 *×* 10^*−*300^). Crucially, the direction of these correlations aligns with the effect sizes (*β*) observed in our ProjWAS. For instance, elevated creatinine is a clinical indicator of kidney dysfunction [29]; the rs2020900 variant is protective against renal failure, which is consistently reflected by its negative correlation with creatinine levels. Similarly, prolonged QRS duration indicates cardiac conduction abnormalities [30]; the rs6795970 variant increases the risk for atrioventricular and left bundle branch blocks, mirroring its positive correlation with the QRS projection phenotype.

Examining the extrema of the projection phenotypes further confirms that they capture the expected morphological and physiological traits (Fig. 3). Consistent with the DISH score [19] and the rs4252548 (*IL11*) projection phenotype being anti-correlated, individuals with the lowest projection scores exhibit pronounced vertebral fusion (indicated by red arrows) in lateral DXA scans. Conversely, individuals with the highest projection scores show normal vertebral anatomy without fusion, corresponding to low DISH scores. This consistency extends to the Fourier-transformed ECG waveforms associated with the rs6795970 variant at the *SCN10A* locus. This projection phenotype is positively correlated with QRS duration, which in turn is associated with conduction disorders [30]. Individuals with the highest projection scores exhibit a wider spread of ECG frequency components indicative of conduction abnormalities, such as bundle branch blocks. In contrast, those with the lowest projection scores present with generally “tighter” frequency spectra. A similar pattern is observed in optical coherence tomography (OCT) imaging, where the projection phenotype for variant rs6420484 at the *TSPAN10* locus is anti-correlated with macular thickness (*ρ* = *−*0.27, *P <* 1 *×* 10^*−*300^). Glaucoma, a condition characterized by progressive thinning of the macula, emerged as a top ProjWAS association for this variant (*β* = 0.979, *P* = 1.28 *×* 10^*−*16^). Evaluating the extremes of the OCT volumes visually corroborates this relationship: high variant projection scores correspond to visibly reduced macular thickness, whereas low scores correspond to preserved macular structure, consistent with prior studies of this gene’s role in macular degeneration risk [31].

**Fig. 3.**
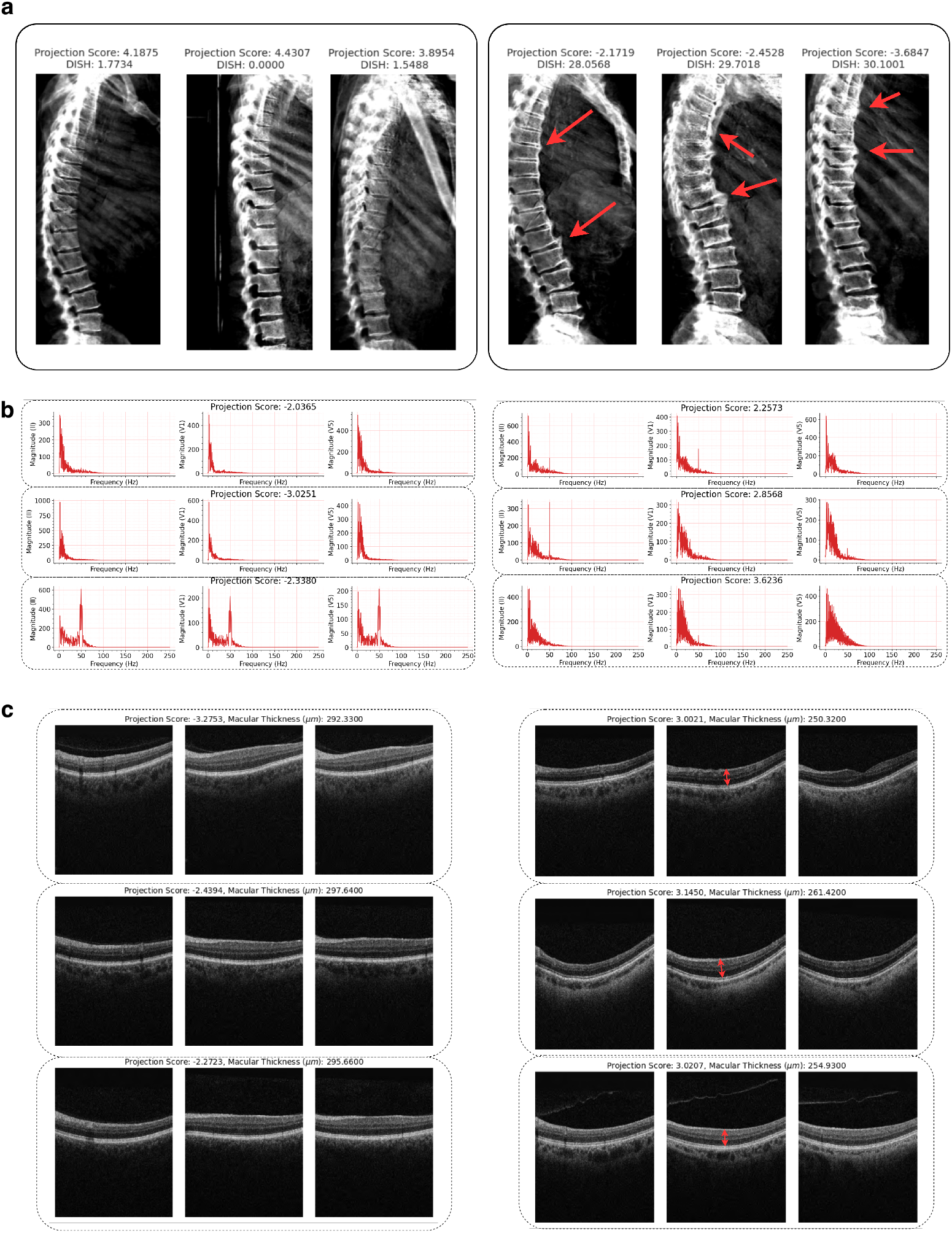
Projection scores reflect structural and electrophysiological variations across diverse modalities. Representative raw imaging and signal data for individuals located at the upper and lower extremes of the calculated projection phenotypes. Across all panels, data displayed on the right represent individuals with observable pathology or altered physiological states. In panels **b** and **c**, each row corresponds to a single individual, showing either three distinct ECG leads or three 3D OCT slices, respectively. **Top (a):** Lateral spine DXA scans associated with the *IL11* locus (rs4252548). Individuals with low projection scores (right) exhibit pronounced vertebral fusion (red arrows), which corresponds to high clinical DISH scores. Conversely, individuals with high projection scores (left) demonstrate normal vertebral anatomy. **Middle (b):** Fast Fourier Transforms (FFTs) of ECG waveforms associated with the *SCN10A* locus (rs6795970). These plots demonstrate observable variations in frequency distributions between the extremes of the projection phenotype, with lower scores (right) exhibiting attenuated high-frequency components compared to the left. **Bottom (c):** Retinal OCT cross-sections associated with the *TSPAN10* locus (rs6420484). High projection scores (right) correlate with visibly reduced macular thickness (red arrows), consistent with increased glaucoma risk, compared to the preserved macular structure seen in individuals with low projection scores (left).

### Projection phenotypes map organ-specific and pleiotropic disease associations

We applied the V2V framework to organ-specific image embeddings derived from abdominal MRI, DXA, brain T1-weighted MRI, cardiac cine MRI, ECG, and OCT data. To interrogate organ-level disease relationships, we defined a set of 9 “primary” modalities that are representative for each of 9 organ systems (eye, brain, heart, lungs, liver, kidney, spleen, pancreas, spine). Leveraging ProjWAS results for each projection phenotype, we aggregated global statistics for loci demonstrating at least one statistically significant disease association. To identify statistically significant disease associations, we used a Bonferroni-corrected threshold [32] adjusting for the number of ICD10 codes tested (811) times the number of independent loci observed in total across these nine modalities using the dominant contrast (2,580), resulting in *P <* 2.4 *×* 10^*−*8^. Specifically, we quantified the number of unique loci per imaging modality associated with at least one phenotype within distinct ICD10 disease categories.

We observe biologically plausible enrichments of specify disease categories in organ-specific contexts (Fig. 4a). For example, OCT-derived loci were predominantly associated with ‘Eye’ diseases compared to other modalities, ‘Genitourinary’ disease associations were highly enriched among kidney-derived loci and brain T1 derived loci with ‘Nervous system’ disease associations. Conversely, associations with ‘Ear’ and ‘Skin’ phenotypes were negligible across all modalities, which is expected given the organs being evaluated. Finally, associations with ‘Circulatory’ and ‘Endocrine / Metabolic’ conditions were captured across most organ systems.

**Fig. 4.**
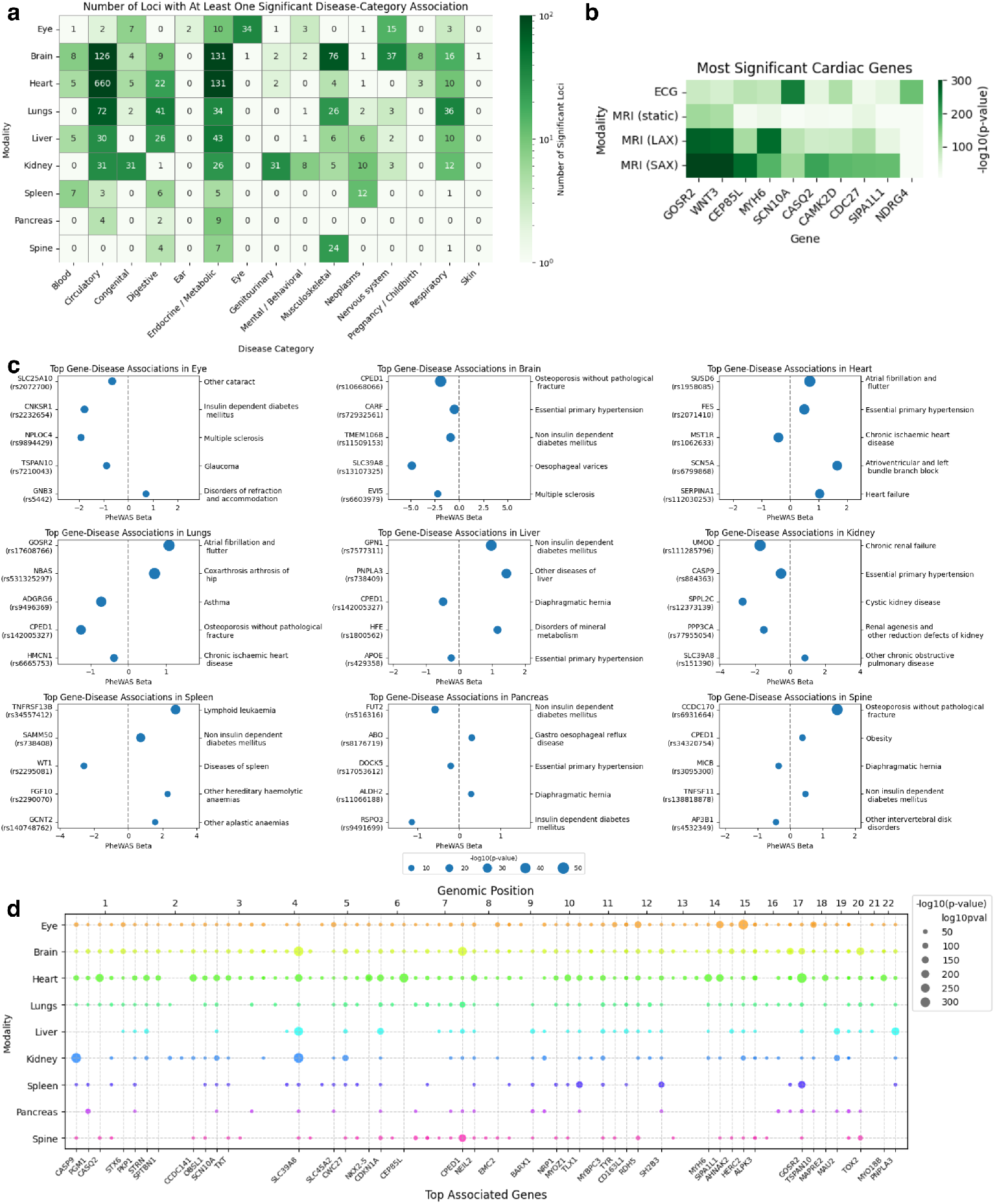
Organ-specific embeddings predict relevant disease categories and show extensive pleiotropy. a, Enrichment of ICD-10 disease categories across projection phenotypes derived from embeddings of nine imaging modalities representing different organ systems. The heatmap displays the count of significant locus-disease associations for a given disease category in a particular organ (using ProjWAS p-values, Bonferroni-corrected for number of independent loci, organs, and diseases tested, *P <* 2.6 *×* 10*−*8). Eye imaging embeddings contain the most associations with eye diseases, brain MRI is most enriched for nervous system, cardiac MRI for circulatory diseases, and so on. Endocrine / metabolic diseases manifest systemically across organs. b, Heatmap of LDA p-values for most significant genes across cardiac modalities: 12-lead ECG, static heart MRI (from abdominal scans), long-axis (LAX) cine MRI and short-axis (SAX) cine MRI. SCN10A (an ion-channel gene involved in cardiac conduction) separates more strongly in ECG than MRI; MYH6 (a muscle contraction gene) separates more strongly in MRI than ECG. c, Top-ranked unique gene-disease associations sorted by ProjWAS significance in imaging modalities from nine organ systems. Point size indicates significance, x-axis represents effect size. We recover well-established disease links, including UMOD with renal failure in kidney, SCN5A with conduction disorders in heart, and CCDC170 with osteoporosis in the spine, confirming that latent representations preserve clinically relevant biological variation. For clarity, we only show the top occurrence of each gene and each disease, removing instances where the same gene or disease occurs multiple times in the top 5. d, Top-down Manhattan plot of most significant gene-modality associations across nine organ systems reveals extensive pleiotropy. Point sizes indicate LDA p-value for lead variant in each genomic region across organs (missing circle indicates non-significant result for this region / organ combination). SH2B3 shows significant associations across most organs (especially spleen and heart), consistent with its broad role in cytokine signaling. SLC39A8 shows broad associations across cardiometabolic and neurological systems.

To assess how different data representations capture distinct biological signals, we compared results from four cardiac modalities acquired from the same subjects: 3D short-axis video MRI, 2D long-axis video MRI, static 3D MRI, and 12-lead ECG Fig. 4b. While many top associations were shared, 3D video MRI yielded the highest number of significant hits, highlighting the value of temporal dynamics over static anatomy. Notably, the method demonstrated modality-specific sensitivity: ECG was uniquely powerful for detecting variants in the ion channel gene SCN10A, involved in conduction velocity [28]), whereas cardiac video MRI showed superior sensitivity for MYH6„ a sarcomeric gene related to contractility [33], highlighting how the projection phenotypes effectively extract the biologically relevant signal specific to the input modality (electrical vs. mechanical).

An analysis of the top unique gene-disease associations, ranked by ProjWAS significance, reveals three findings (Fig. 4c). First, the associated disease phenotypes demonstrate biological concordance with their respective organ embeddings. For example, liver-derived embeddings mapped appropriately to relevant systemic and metabolic conditions, including non-insulin-dependent diabetes mellitus, mineral metabolism disorders, and other diseases of liver. Second, ProjWAS recovers established gene-disease relationships previously documented in the literature. Notable validations include *PNPLA3* (rs738409) with liver disease [34], *UMOD* (rs111285796) with chronic renal failure [35], *SCN5A* (rs6799868) with left bundle branch block [36], and *CCDC170* (rs6931664) with osteoporosis [37]. Finally, the directionality of the ProjWAS effect sizes (*β*) reveals the protective or risk effects of these alleles. For example, the variant rs111285796 in *UMOD* exhibits a negative effect size on chronic renal failure, concordant with previously reported associations with kidney function [38]. Conversely, positive effect sizes correctly indicate increased risk, as observed with rs6931664 in *CCDC170* increasing the risk of osteoporosis, aligning with previous studies of bone mineral density [5].

By comparing top associations across modalities in Fig. 4d, we observed pleiotropic roles of multiple genes. For instance, the projection axis optimized for rs3184504 in SH2B3—a key regulator of cytokine signaling and hematopoiesis—was highly significant in spleen embeddings, with moderate significance in brain, heart, aorta, liver, and pancreas embeddings. This mirrors its known involvement in diverse pathologies ranging from hypertension to myeloproliferative disorders [39]. Similarly, projection of variants in SLC39A8 displayed strong associations across brain, heart, kidney, and liver, consistent with its role as a transporter of manganese and zinc, essential for neurological and cardiometabolic function [40].

### V2V replicates GWAS associations while capturing additional signal

We compared the results of our method (dominant contrast) to GWAS on the same cohort using four categories of quantitative traits - cardiac IDPs (61 traits derived from short-axis cardiac MRI [41]), brain IDPs (1436 traits derived from T1-weighted MRI [11]), NMR metabolomics (251 traits corresponding to metabolite abundance [17]) and PPP proteomics (2913 traits corresponding to protein abundance [16]). For each category, we aggregated the GWAS results (picking the best p-value across all traits for a given variant, with Bonferroni correction) and compared to the analogous modality in our V2V results. We then identified variants that are significant in (1) GWAS-only, (2) V2V-only, or (3) both V2V and GWAS at different significance thresholds, along with the overall correlations of the p-values (Fig. 5).

**Fig. 5.**
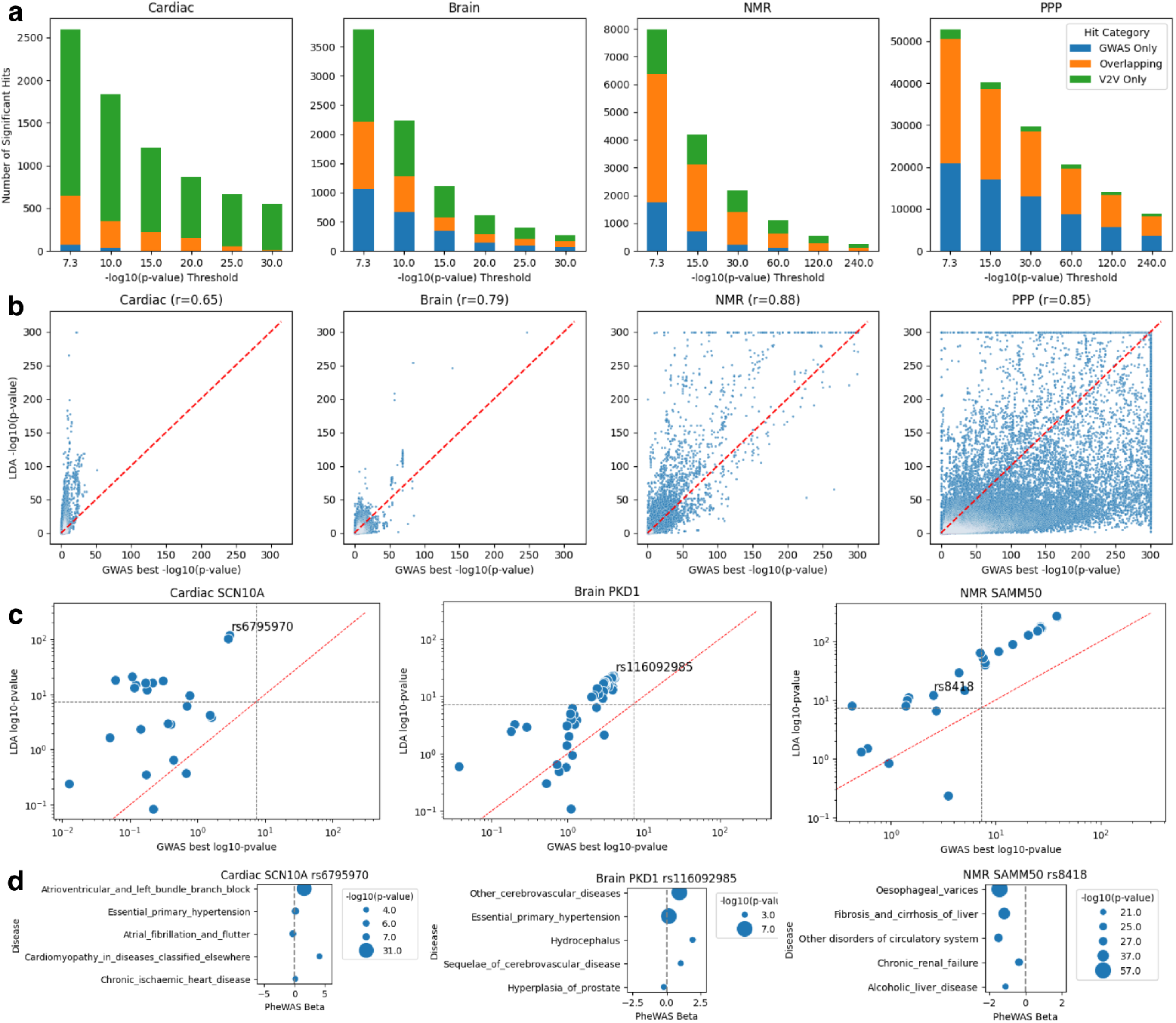
Comparison to Cohort-Matched GWAS on Engineered Phenotypes. a, Comparison of significant variants found by our method vs GWAS on panels of modality-specific phenotypes for cardiac, brain, NMR and PPP. GWAS results are aggregated using Bonferroni-corrected min-p-value. At each significance threshold, we compare number of significant variants found in V2V-only (green), GWAS-only (blue), or both (orange). Using cardiac SAX embeddings, we recapture almost all variants founds from ventricular cardiac IDPs, along with many additional ones. Using brain anatomical T1 embeddings, we capture the majority of variants found from GWAS of T1 brain IDPs. In NMR, our method uses the same metabolite features as GWAS, and there is correspondingly more overlap in the results. In PPP, we use the same protein features as GWAS but see less favorable performance, owing to the highly univariate relationship between cis-pQTLs and protein abundance. b, Scatter plot showing correlation of log10-pvalues (capped at 300) from our method vs best GWAS result for the four modalities. c, Gene-specific scatter plots showing p-value correlations for 3 genes with known modality-specific disease effects where V2V shows greater sensitivity than GWAS: SCN10A in cardiac, PKD1 in brain, and SAMM50 in NMR. Grey dotted lines indicate genome-wide significance we highlight variants that are significant in V2V but not GWAS. d, Top ProjWAS results for highlighted variants from panel c that are significant in V2V but not GWAS, illustrating relevant disease associations (note that esophageal varices are typically caused by liver cirrhosis, supporting the known SAMM50 liver disease link, captured here purely from metabolomics).

For the cardiac and brain comparisons, we compared GWAS on engineered features to V2V on image embeddings. For example, the cardiac IDPs consist of 61 features derived from short-axis video (e.g. ventricular volumes and ejection fraction) whereas the cardiac embeddings consist of 768 feature dimensions from the same short-axis video. We chose this comparison to illustrate the phenotype-free nature of our method. Conversely, for the metabolomics and proteomics comparisons, we directly used the same set of features for both GWAS and V2V, providing a strong baseline.

Our results suggest that V2V can replicate many or most of the hits obtained from GWAS, depending on the modality, while also capturing some “V2V-novel” hits which are not significant in the matched GWAS. We also see generally strong correlation between the GWAS and V2V p-values, with mildly significant variants in GWAS getting “boosted” to higher significance in V2V. This effect is most pronounced in cardiac MRI, owing to the relatively small number of engineered phenotypes. For brain, the results are somewhat less favorable, but there are still more V2V-only hits than GWAS-only hits. In NMR, there is relatively more overlap between V2V and GWAS, since we are using the exact same features, but V2V and GWAS each capture some hits that the other method does not. In the PPP, we see the smallest fraction of hits replicated in V2V relative to GWAS, although the majority of GWAS hits are also captured by V2V, with only a small fraction of V2V-novel hits. This aligns with the intuition that PPP features are already highly specific, and in particular, we expect strong associations of individual variants in a gene with the abundance of its corresponding protein in plasma (cis-pQTLs). Therefore, GWAS of individual protein abundances is a difficult baseline to beat, especially since V2V is not well-suited to selecting individual sparse proteins / features when learning a classifier.

We investigated the top V2V-novel hits to look for interesting biology. In cardiac MRI, we identified several variants in SCN10A (e.g. rs6795970 missense) which has established associations with cardiac conduction and arrhythmias [3, 20], which were highly significant in V2V but not in the GWAS of cardiac IDPs. In brain MRI, we observed several novel associations with variants in PKD1 (rs116092985 missense), most commonly linked to polycystic kidney disease [42]. PKD1 is highly expressed in the brain and blood and and variants have been linked to aneurysms and other cerebrovascular conditions linked to PKD-induced hypertension [43]. Correspondingly, we found that our top ProjWAS hits for these variants were cerebrovascular disease and hypertension, strengthening the connection with prior evidence. In NMR, we found several variants in SAMM50 and PNPLA3 (rs8418 missense, rs2294918 missense) which were not significant in the matched GWAS, with “fibrosis and cirrhosis of liver” as one of the top ProjWAS hits, matching the well-established role of this gene in NAFLD [34, 44]. These were previously identified as borderline significant in GWAS of hepatic fat from CT imaging [45], but not previously detected from metabolic markers.

### Novel associations include convergent signatures across organs

Comparing our results across nine organ systems, we identified 474 novel organ-locus associations not found in GWAS Catalog [15]. We defined “novel” loci in the context of a specific organ by grouping GWAS Catalog traits into organ-specific categories based on the Experimental Factor Ontology (EFO) [46]. For example, to identify “brain-novel” loci, we compare to any GWAS Catalog trait that maps to brain disease or brain-derived attributes in the EFO.

We identified variants in the LRRIQ1 gene that show significant separation in kidney, liver, heart, and lung embeddings, centered on rs758375774, a frameshift variant with predicted loss-of-function (LoF) effect (Fig. 6). While LRRIQ1 is recognized for its role in sperm production [23], previous association studies point to disparate traits like neuroimaging features [47] and facial morphology [48], with no consistent disease associations. Our method revealed novel organ-specific genetic separations with a consistent disease signature that has not been previously reported. In kidney, the projection phenotype for this LoF variant was strongly associated with cystic kidney disease and hypertension. In liver, it’s associated with atrial fibrillation (AFib) and hypertension. In heart, associations were found with AFib, chronic ischemic heart disease and acute myocardial infarction (MI). Notably, the ProjWAS effect sizes for hypertension, AFib and ischemic heart disease were negative, suggesting that this LoF variant may exert a protective effect against cardiovascular decline. The consistency of this signal across multiple independent organ systems provides additional support and suggests that LRRIQ1 plays a broader role in systemic vascular health than previously understood, perhaps being an example of antagonistic pleiotropy given its role in fertility.

**Fig. 6.**
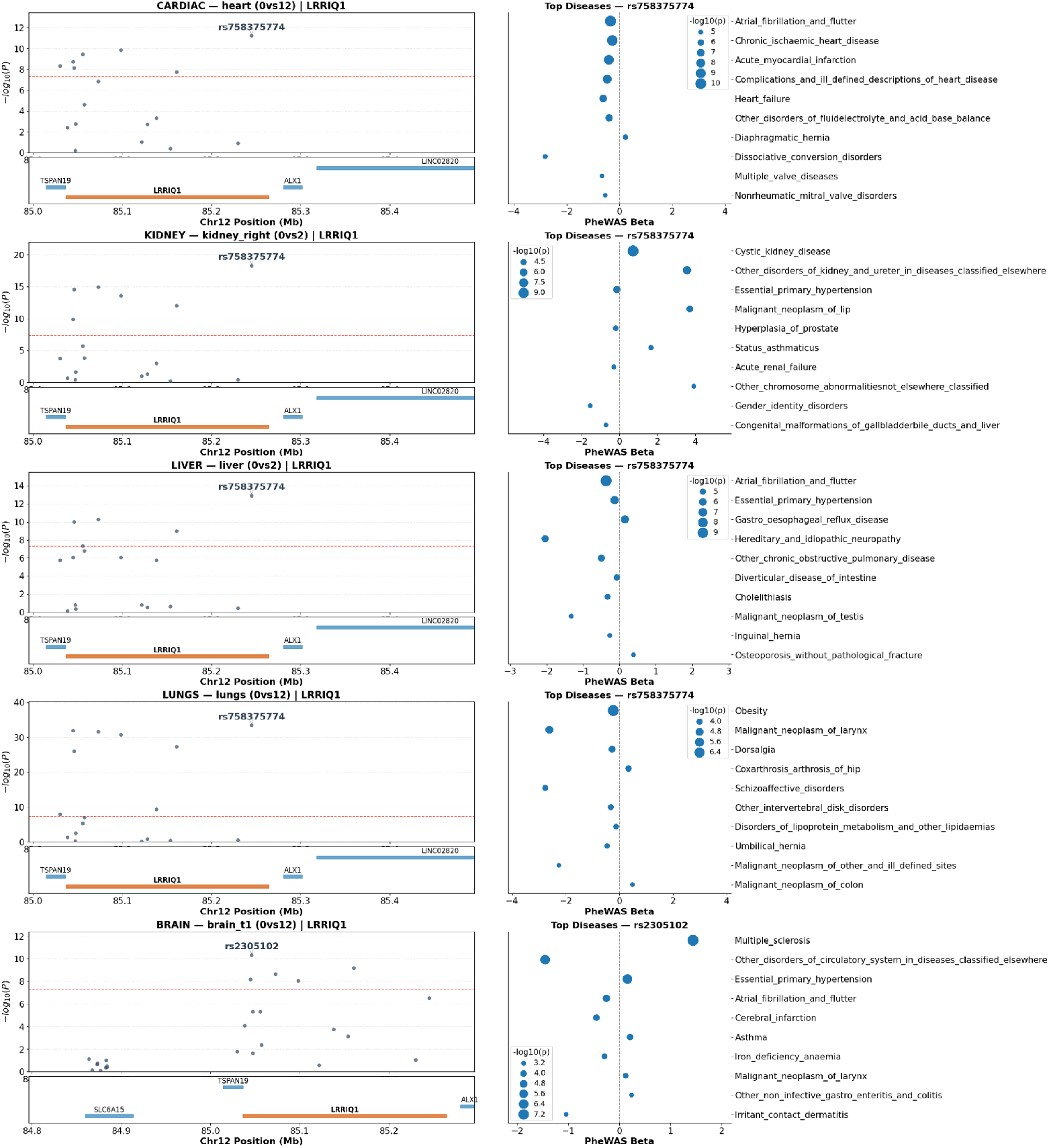
Discovery of Novel Organ-Disease Associations in LRRIQ1. (left column) Locus- Zoom plots of lead variants discovered in LRRIQ1 across five organ systems - cardiac, kidney, liver, lungs, brain. Note that the first four organs share the same lead variant, a frameshift mutation. (right column) corresponding top ProjWAS results for the lead variant in each organ, showing some consistent disease signatures across organs (AFib in heart, liver, brain; hypertension in kidney, liver, brain) as well as noteworthy organ-specific effects (cystic kidney disease, multiple sclerosis). The hypertension and AFib effects are protective in the case of the frameshift variant. GWAS Catalog does not report any prior associations with heart, kidney, or liver-specific traits.

Our framework also highlighted two missense variants in LRRC37A2 (rs1863115, rs199913382) as having pleiotropic effects (Figure S4). While LRRC37A2 has been previously implicated in Parkinson’s disease [49] and longevity [50], its connection to metabolic health has remained speculative. Our method approach identified significant separation of this variant across the brain, heart, kidney, lung, and spleen embeddings. Significant associations with Type 2 Diabetes (T2D) were detected across the spleen and brain, hypertension across brain / heart / lungs, and AFib across heart and lungs. The separation in spleen is particularly significant (*log*10(*p*) *>* 150) and novel compared to GWAS Catalog, with the diabetes and heart disease associations suggesting an interaction with cardiometabolic function.

In retinal OCT imaging, we identified a cluster of variants in a multi-gene region spanning ZNF593OS / CNKSR1 / TRIM63 / CATSPER4 that consistently associate with Type 1 Diabetes (T1D), glaucoma, and multiple sclerosis (MS) (Figures S5, S6). Notably, variants in CATSPER4 and CNKSR1 exhibit a protective signature across T1D, T2D, and MS based on our ProjWAS. In heart imaging, the projections of nearby variants are linked to conduction disorders, AFib and angina. These loci are novel in terms of their association with eye measurements. The ProjWAS results recapitulate established links between diabetes / glaucoma [51] and MS / optic neuritis [52]; however, the convergence of associations between diabetes and MS is an under-explored area.

Beyond these initial examples, our framework identified a wide range of other gene-organ associations that have little or no prior support in GWAS Catalog, but have suggestive disease-ProjWAS results in organ-specific contexts, opening avenues for further investigation (Figures S7-S11). In the kidney, we found that the metabolic enzyme PCK2 is significantly associated with chronic renal failure and cystic kidney disease. This could reflect a disruption of renal energy metabolism or redox balance, as PCK2 is increasingly recognized as a factor in renal interstitial fibrosis [53]. In the brain, the fibrosis-related gene ITGA11 emerged as a novel factor in cerebrovascular health, showing a specific association with vascular syndromes which has some support [54]. We also identified multiple cardiac disease associations the SUPT3H/RUNX2 locus; while its link to bone formation is established [55], our associations with atrial fibrillation and hypertension suggests that vascular calcification may be driven by shared osteogenic mechanisms, which is an active research area [56]. The ubiquitin-conjugating enzyme UBE2U was highlighted as a potential novel driver of heart disease, consistent with emerging evidence linking ubiquitin-proteasome system (UPS) dysfunction to heart failure [57]. The ubiquitin-specific peptidase USP37 was strongly linked to Alzheimer’s disease in the brain and cardiac diseases in the heart, with not much prior literature, although the locus is being actively studied for its role in glycogen aggregation in astrocytes with potential links to neurodegeneration [58].

### Multivariate framework disentangles regulatory interactions masked in univariate Omics analysis

Our V2V framework is not limited to embeddings - it can be applied to any set of continuous measurements, such as a panel of blood biomarkers. To demonstrate this, we applied the method to high-dimensional proteomic (PPP, 2913 proteins) and metabolomic (NMR, 251 metabolites) panels. Unlike univariate GWAS, which tests one protein / metabolite at a time [16], our linear discriminant approach learns multivariate effects, assigning weights to markers that maximize genotypic separation. Notably, V2V successfully replicated several well established gene-disease associations. In the PPP data, these included the APOE rs429358 variant with Alzheimer’s disease [59], MMP3 with aortic aneurysm and dissection [60], and ACE with essential primary hypertension [61] (Fig. 7a). Similarly, in the NMR data, we identified expected associations, such as FAM47E with chronic renal failure and APOB with disorders of lipoprotein metabolism.

**Fig. 7.**
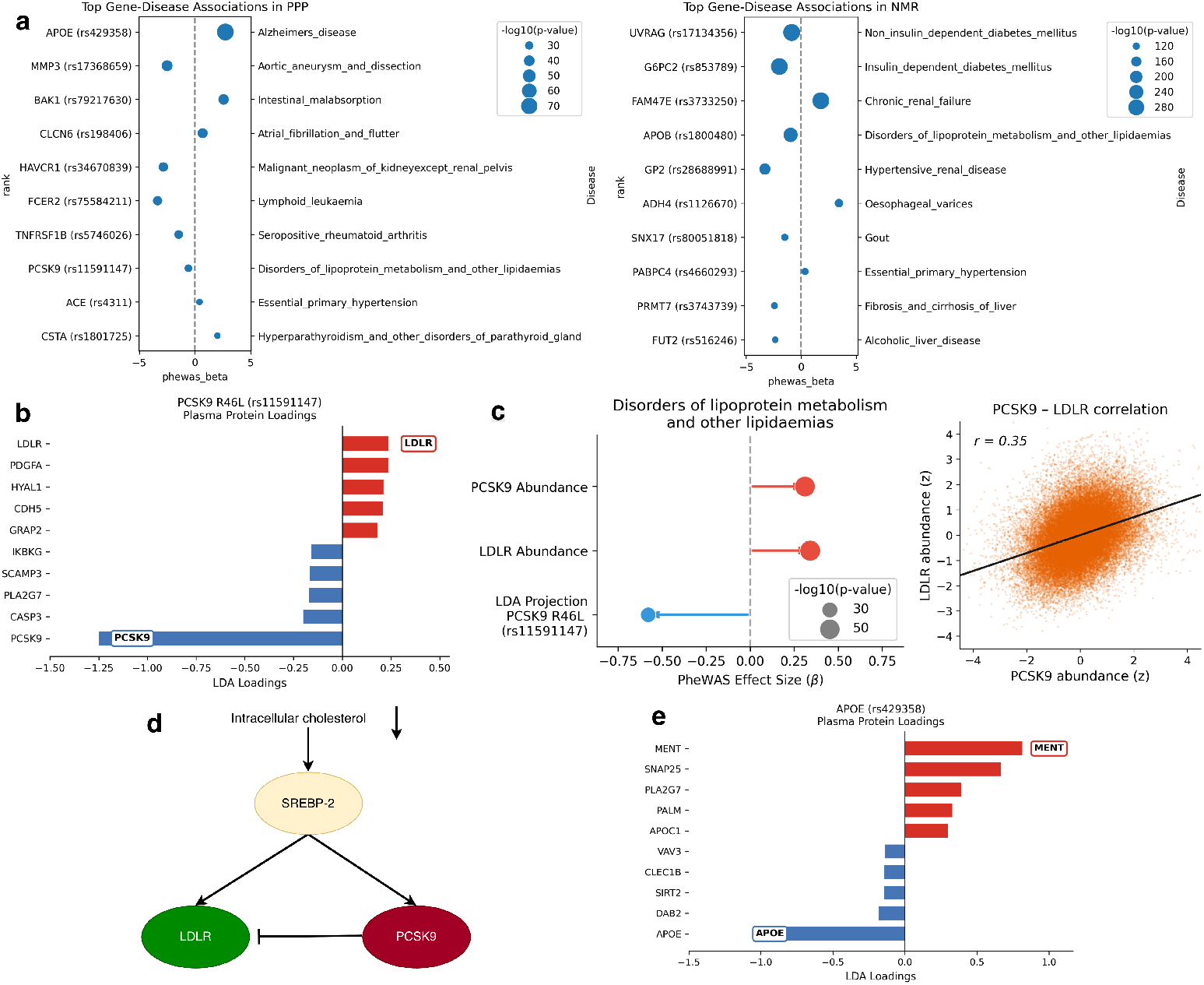
Disentangling multivariate mechanisms in high-dimensional omics. a, Top gene- disease ProjWAS associations identified in PPP proteomics (left) and NMR metabolomics (right), capturing known associations like APOE-Alzheimer’s, PCSK9-hypercholesterolemia, and FAM47E- renal failure. Genes in the HLA region are excluded for clarity. b, Feature weights of the linear classifier for the PCSK9 rs11591147 (R46L) loss-of-function variant. The model assigns a negative weight to PCSK9 protein abundance and a positive weight to LDLR abundance, reflecting the known mechanism of PCSK9 inhibition leading to decreased LDLR degradation. c, Comparison to univariate analysis of PCSK9-LDLR, showing a positive association of both PCSK9 and LDLR protein abundances with hypercholesterolemia (left), and a positive correlation of PCSK9 with LDLR (right). d, Schematic of the PCSK9-LDLR interaction pathway illustrating upstream co-regulation of both PCSK9 and LDLR (e.g., SREBP-2). e, Feature weights for APOE rs429358 (part of the APOE-*e*4 genotype), showing negative association with APOE and positive association with MENT, a protein that has previously been positively associated with *e*4 carrier status in individuals with Alzheimer’s disease.

For each variant, we compute not only a projection score but also the underlying LDA projection weights (loadings). When utilizing measured biomarkers like PPP and NMR as input vectors, these loadings directly measure the contribution of each biomarker to the projection score, allowing us to interpet the score in terms of upregulation / downregulation. For instance, examining the LDA protein loadings for the APOE rs429358 variant reveals that the most negative weight is assigned to APOE protein abundance, while the most positive weight is assigned to the MENT protein (Fig. 7e). This independently replicates previous findings demonstrating that MENT (C1orf56) is upregulated in APOE-_*∈*_4 individuals with Alzheimer’s disease [62].

Furthermore, the multivariate nature of the V2V framework inherently captures complex interactions between variables that univariate methods miss. To demonstrate this, we investigated the LDA weights for PCSK9 variant rs11591147, which yielded *Disorders of lipoprotein metabolism and other lipidaemias* (the ICD10 category which broadly maps to high cholesterol) as its top ProjWAS association in the PPP dataset. This variant, commonly called R46L, is a known loss-of-function variant; reduced levels of functional PCSK9 lead to decreased degradation of the low-density lipoprotein receptor (LDLR), thereby increasing LDLR abundance [63]. Our LDA loadings recapitulate this biology, assigning large opposing weights to PCSK9 (negative) and LDLR (positive). Crucially, standard univariate approaches fail to capture this regulatory axis; because PCSK9 and LDLR are co-regulated by SREBP-2 in a feedback loop, univariate analyses are prone to confounding and report positive ProjWAS betas for both proteins in relation to lipid disorders. By focusing instead on the axis of genetic separation, our method disentangles the variant effect from compensatory protein expression, yielding a negative beta that reflects the variant’s protective effect.

## Discussion

Vector2Variant demonstrates that it is possible to discover both well-validated and novel relationships between genetics, organ physiology, and disease without manual curation of phenotypic traits. This approach offers a systematic alternative to traditional GWAS, which frequently rely on the reduction of dense biological data into a small number of predefined, handcrafted features. By identifying the specific axes in multivariate space along which genotype classes are most separable, V2V derives “projection phenotypes” that reflect the structure of the underlying data rather than a researcher’s a priori assumptions. Our findings across diverse organs and modalities indicate that these learned axes are biologically coherent, as evidenced by their correlation with established clinical biomarkers and their ability to recover known gene-disease relationships like CASP9 and SCN10A alongside pleiotropic effects. Furthermore, the detection of novel signals, such as the LRRIQ1 cardiovascular association, suggests that this framework can complement existing discovery pipelines by surfacing multivariate patterns that might be obscured in standard univariate analyses. While the method’s performance is inherently linked to the quality of the input embeddings, the consistency of results across diverse organ systems suggests that a variant-centric model is a viable strategy for navigating the high-dimensional data landscape of modern biobanks.

Several multivariate GWAS methods have been proposed, each addressing a different facet of the phenotype dimensionality problem. PCA-GWAS [11] frameworks such as REGLE [64] project phenotypes onto axes of maximum variance, but these axes are not optimized for genetic separation and may fail to capture biologically relevant variation that is distributed across many components; we provide examples where known variant-phenotype associations are not recoverable from the top principal components of the same embeddings despite these components explaining the majority of total variance (Supplement S4). MOSTest [65] aggregates univariate statistics across phenotypes into a powerful omnibus test, but yields no interpretable effect direction or per-individual scores. MultiPhen [66] tests whether phenotypes jointly predict genotype, but depends on asymptotic calibration assumptions that break down when the number of features rivals the number of variant carriers (Supplement S1). Many methods require predefined scalar phenotypes. Our framework sidesteps these limitations by learning a variant-specific discriminative axis, producing scores that can be interpreted with downstream ProjWAS, and maintaining calibrated statistics even in severely high-dimensional settings through out-of-sample evaluation (Supplement S1). To our knowledge, this represents one of the first comprehensive studies using self-supervised neural network embeddings to directly drive genome-wide genetic discovery, without an intermediate feature selection step, alleviating the need for domain-specific phenotype engineering which has traditionally been a barrier to the unbiased interrogation of complex data modalities.

Our approach draws on the same core insight as multi-voxel pattern analysis (MVPA) in functional neuroimaging [67]. MVPA demonstrated that cognitive states could be decoded from distributed spatial patterns across voxels that were individually non-informative, a finding that reshaped how the field interpreted brain imaging data. Vector2Variant applies this insight to genetics: just as MVPA trains classifiers to find the neural pattern that best separates experimental conditions, our method trains a linear discriminant model to find the embedding pattern that best separates genotype classes. In both cases, the key departure is shifting from mass univariate testing of individual features to learning a single multivariate axis optimized for discrimination. This represents an inversion of the traditional GWAS question: rather than asking which variants associate with a given phenotype, the method asks what phenotype a given variant *defines* inside the embedding. This inversion is well-suited to the current state of human genetics, where the cost of genotyping has fallen dramatically while phenotyping, particularly from dense modalities requiring domain expertise, remains the primary bottleneck. The PCSK9 result illustrates why the multivariate formulation matters in practice - the individual protein abundances of PCSK9 and LDLR are both positively correlated with hypercholesterolemia when tested in a univariate fashion, yet the discriminative axis grounded on a known LoF variant correctly recovers their inverse mechanistic relationship, much as an MVPA classifier can recover a neural pattern that no single voxel expresses.

From a translational perspective, the ProjWAS effect directions, when paired with variant annotations (e.g., loss-of-function, missense), provide initial evidence for whether perturbation of gene function is protective or deleterious for a given disease. A researcher investigating an association of interest can further refine interpretation by extending ProjWAS beyond ICD-10 codes to include domain-specific phenotypes, biomarkers, or quantitative traits relevant to the association. More broadly, there is growing evidence that drug targets with human genetic support succeed at higher rates in clinical trials [68, 69]. V2V expands the genetic evidence base by surfacing associations that the reliance on predefined phenotypes has traditionally prevented from being discovered. The projection phenotype need not be treated as an endpoint; rather, it serves as a starting point for further genetic and functional investigation, now applied to phenotypic variation that was previously inaccessible.

The statistical power of V2V is governed by the same factors as GWAS, primarily minor allele frequency and cohort size, and the method is powered in the same MAF regime. We empirically observe sensitivity to rare coding variants below 1% MAF, including ClinVar pathogenic alleles such as *TNFRSF13B* rs34557412 (MAF ≈ 0.7%) and *CSRP3* rs45550635 (MAF ≈ 0.5%), with biologically concordant ProjWAS results (Supplement S2). However, for very rare variants where carrier counts are insufficient for robust classifier training, the per-variant formulation becomes a limitation. A natural extension is gene-level aggregation, combining carriers of any qualifying rare variant within a gene into a single class for LDA training to increase the effective sample size. Analogous to gene burden approaches, this assumes that qualifying variants shift the embedding in a consistent direction. We leave this extension for future work. As with any association framework, V2V operates within constraints that shape the interpretation of its results. The method can only recover biology that the input representations encode; if a self-supervised model discards a biologically relevant feature during training, V2V cannot detect its genetic basis. However, accumulating evidence suggests that modern self-supervised representations retain far more clinically relevant information than handcrafted features typically extract [8–10]. In our own data, projection scores correlate with orthogonal biomarkers (serum creatinine, liver iron, QRS duration) that were never provided to the embedding models, and this information content will only grow as foundation models improve. A related concern is the linearity of the discriminant model - LDA identifies only linear axes of separation, and nonlinear effects would be missed. In practice, this constraint may be less restrictive than it appears, because deep neural networks already capture complex nonlinear relationships in their learned representations, and there is substantial evidence that the resulting embedding spaces tend to encode semantic factors linearly [7, 70]. Several additional choices shape the boundaries of the present study. All analyses were restricted to participants of European genetic ancestry, and the transferability of both embedding models and projection phenotypes to other populations remains untested.

The framework tests one variant at a time, and epistatic interactions or gene-gene effects are not modeled. While ProjWAS associations provide interpretable disease links for each projection phenotype, these remain statistical correlations; establishing causality requires dedicated follow-up through causal inference and functional genomics approaches. Because V2V learns a separate projection axis for each variant, it does not produce a single shared phenotype amenable to standard polygenic analyses. Thus, downstream applications that rely on the additive polygenic model, such as polygenic risk score construction, heritability estimation, and cross-variant aggregation, are not directly supported by the current framework. We leave the integration of fine-mapping, colocalization, and Mendelian randomization with projection phenotypes for future work.

Looking ahead, the framework’s modality-agnostic design positions it to benefit directly from ongoing advances in machine learning. Another direction is cross-modal integration. Currently, each modality is analyzed independently, but contrastive learning objectives analogous to those used in vision-language models [71] could enforce representational consistency across organs, learning a shared embedding space in which the same variant’s effect is aligned across modalities. In addition, such architectures could bridge imaging representations directly to structured clinical data such as electronic health records (EHR), linking the rich but uninterpretable biological variation captured by self-supervised models to the interpretable but sparse phenotypic information in clinical databases, within a single framework.

Our method demonstrates that the combination of self-supervised representations and a simple discriminative model is sufficient to convert high-dimensional biological measurements into genetic discoveries and testable clinical hypotheses, without requiring handcrafted phenotypes. As biobanks grow in scale, diversity, and modality coverage, frameworks that bypass the phenotyping bottleneck will become increasingly valuable for genetic discovery.

## Methods

### Notations

Let 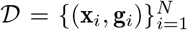 denote a dataset of *N* individuals. Let x_*i*_ *∈* ℝ^*D*^ represent the *D*-dimensional feature embedding for individual *i*, derived from a high-dimensional modality via a representation learning function *f*: *ℐ →* ℝ^*D*^. Let g_*i*_ ∈ { 0, 1, 2} ^*V*^ represent the genotype vector for *V* variants, where entries correspond to the count of alternate alleles (0 for homozygous reference, 1 for heterozygous, and 2 for homozygous alternate). We define *𝒳*∈ ℝ^*N ×D*^ as the full embedding matrix and 𝒢 ∈ {0, 1, 2} ^*N×V*^ as the full genotype matrix. For a specific variant *v*, we define two distinct classes of individuals:

1. **Homozygous Contrast (Primary):** *C*_0_ = {*i* |*g*_*i,v*_ = 0} and *C*_1_ = {*i* |*g*_*i,v*_ = 2}. This considers the genotypic extremes and ignores heterozygous population.
2. **Dominant Contrast (Secondary):** *C*_0_ = *{i* | *g*_*i,v*_ = 0*}* and *C*_1_ = *{i* | *g*_*i,v*_ *∈* {1, 2}}. This combines heterozygous with homozygous alternates.

### Covariate Adjustment

To ensure that learned projections capture genetic effects rather than confounding demographic structure, we perform covariate adjustment prior to running our proposed method. Let C∈ ℝ^*N ×K*^ be the matrix of *K* covariates (including age, sex, genotyping array, imaging center, and BMI). Categorical variables with *L* classes were encoded as *L* one-hot vectors. We first fit a linear model to predict the embeddings from covariates by solving the least-squares optimization problem:

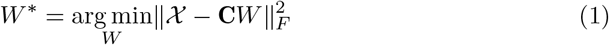

We then compute the residualized embeddings 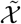 by subtracting the covariate-explained component:

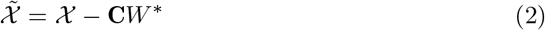

All subsequent operations are performed on the residualized feature matrix 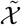.

### Linear Discriminant Analysis (LDA)

Many phenotypes of interest (e.g., volume, iron concentration) can be predicted from the embeddings using linear regression (as shown in Fig. 2), which could be integrated into a traditional GWAS pipeline to identify genetic associations. However, our proposed method bypasses this explicit linear regression step to directly obtain these genetic associations.

Suppose there exists an unobserved continuous latent phenotype *p* that is perfectly captured by a linear projection of the true latent features, such that 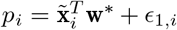 for some ideal projection vector w^*\**^. If this latent phenotype is significantly associated with a genetic variant *v*, it can be modeled as 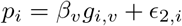, where the effect size is non-zero (*β*_*v*_ ≠ 0).

By evaluating the expected phenotype for the *C*_0_ and *C*_1_ groups, we can link the genotypic effect back to the embedding space. Assuming centered noise and focusing on the homozygous contrast (*g*_*i,v*_ ∈ {0, 2}), the expected phenotypes based on the projection are:

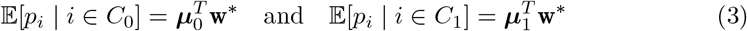

Concurrently, the expected phenotypes based on the genetic association model are:

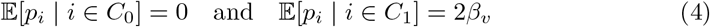

Equating the differences between the two group formulations yields (***µ***_1_*−****µ***_0_)^*T*^ w^*\**^= 2*β*_*v*_. Because the genetic association is significant (*β*_*v*_ 0), the inner product must be non-zero, which strictly dictates that the mean embeddings of the two genetic classes must be distinct:

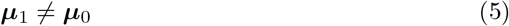

Motivated by this observation, we seek the projection axis w that maximizes the Fisher criterion [25]:

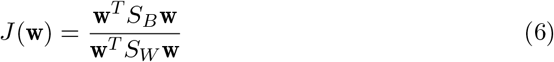

where *S*_*B*_ = (***µ***_1_*−* ***µ***_0_)(***µ***_1_*−* ***µ***_0_)^*T*^ is the between-class scatter matrix and *S*_*W*_ is the within-class scatter matrix, defined as *S*_*W*_ = Σ_0_ + Σ_1_ (with Σ_*k*_ representing the unscaled covariance matrix of class *k*). In the binary classification setting, *S*_*B*_ has rank 1, and the transformation *S*_*B*_w is always proportional to the mean difference vector (***µ***_1_*−****µ***_0_). Consequently, the optimal projection vector 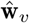 that maximizes *J* (w) simplifies analytically to:

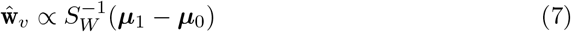

In high-dimensional settings where the embedding dimension *D* approaches or exceeds the number of samples in the minority class (*D »*|*C*_1_|), the empirical within-class scatter matrix *S*_*W*_ can become singular or ill-conditioned. To address this, we replace *S*_*W*_ with the Ledoit-Wolf shrinkage estimator *Ŝ* _*LW*_ [72]. This estimator constructs a well-conditioned covariance matrix by finding the optimal convex combination of the empirical sample covariance *Ŝ*_emp_ and a low-variance target matrix (the identity matrix *I* scaled by the average eigenvalue *v*):

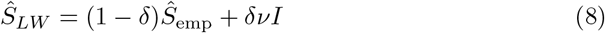

The shrinkage coefficient *δ* ∈ [0, 1] is analytically calculated to minimize the Mean Squared Error (MSE) between the estimated and true population covariance matrices. The final robust projection axis is computed as 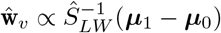.

Because the solution to the Fisher criterion is scale-invariant and its sign is mathematically arbitrary, we impose a strict orientation constraint on the projection axis 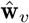. By ensuring that the projected mean of the carrier class is greater than that of the reference class 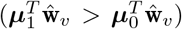, the sign of downstream ProjWAS associations unambiguously indicates whether the alternate allele drives a protective benefit or an increased disease risk.

While our proposed framework utilizes LDA, any suitable linear model could theoretically be applied to identify the projection axis. We selected LDA because empirical evaluations demonstrated superior performance over other linear models in this context, alongside advantages for scalability. The exploration and benchmarking of alternative linear models are left for future work.

### Cross-Validation and Statistical Inference

To mitigate overfitting in high-dimensional settings where the feature count often exceeds the number of rare variant carriers, we employ a rigorous stratified cross-validation scheme. For each variant *v*, the available cohort is partitioned into a training set 𝒯 (40%) and a validation set 𝒱 (60%), stratified by the number of alternate alleles to ensure adequate representation of carriers in both sets.

The projection vector 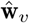 is estimated solely using the training set 𝒯. We then project the held-out validation set 𝒱 onto this axis to generate continuous phenotype scores:

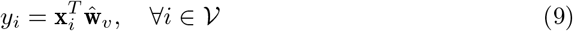

Statistical significance is not assessed by the separation of the classes used for training. Instead, we evaluate whether the projected phenotype score scales additively with allele dosage across the entire validation population. We fit a linear regression model on the validation scores:

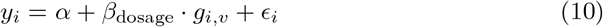

where *g*_*i,v*_ ∈ {0, 1, 2} represents the allele dosage. The significance of the variant-phenotype association is quantified by the p-value of the coefficient *β*_dosage_. This approach validates that the learned axis captures a continuous phenotypic gradient (Reference *<* Heterozygous *<* Homozygous Alternate) rather than merely separating the extremes used during training. To ensure robustness against random partitioning variability, we repeat this procedure over *k* = 3 random splits and compute the final significance as the harmonic mean of the resulting p-values [73].

### Projection-Phenome Association Study (ProjWAS)

For variants passing the genome-wide significance threshold (*P <* 5*×*10^*−*8^), we utilize the continuous scores y to uncover the broader phenotypic correlates of the genetic variation. While the LDA projection identifies *if* a variant is associated with a shift in the high-dimensional representation, the ProjWAS step elucidates *how* this shifted profile relates to clinical outcomes. We treat the projection score as a quantitative trait and test for associations across the full spectrum of medical diagnoses.

We utilize ICD-10 hospital records (UKB field 41270) to define distinct case-control cohorts, following the protocol established by the `PHEASANT` library [74] and grouping ICD-10 codes by category (3-character prefix). To ensure robustness, we restrict our analysis to diseases with at least 5 cases in the cohort. This results in a median of *M* = 811 distinct phenotypes tested (varying slightly by data modality). For each disease *d*, we fit a logistic regression model:

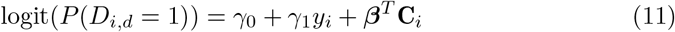

where *D*_*i,d*_ is the binary status of disease *d* for individual *i*. To control for confounding, we include a vector of covariates C_*i*_ (age, sex, and principal components of ancestry) directly in the model. To ensure our clinical associations are robust to the random partitioning used during LDA training, we repeat the ProjWAS procedure across all *k* = 3 cross-validation folds. For each fold, we project the entire cohort (training + validation) onto the learned axis to compute scores, run the association test, and finally aggregate the resulting p-values using the harmonic mean. Significant associations in this step provide clinically interpretable labels for the projection axes discovered in the embedding space.

### Scalable Computational Framework

Executing high-dimensional association scans and subsequent phenome-wide analyses at genome-wide scale equires substantial computational resources. To achieve feasible runtimes, we implemented a highly parallelized architecture tailored to the specific computational bottlenecks of each pipeline stage.

#### GPU-Accelerated High-Dimensional Associations

For the primary variant screening, we implemented a custom, GPU-accelerated version of the Ledoit-Wolf LDA algorithm in `pytorch`. This implementation vectorizes the empirical covariance matrix estimation and eigenvalue shrinkage, enabling the simultaneous optimization of projection axes across thousands of variants. The implementation natively manages the variant-specific training and validation partitions required for our cross-validation scheme directly in GPU memory. This approach scales horizontally across an arbitrary number of hardware accelerators, massively reducing the compute time required for the genome-wide scans.

#### CPU-Distributed Projection-Phenome Associations (ProjWAS)

The downstream ProjWAS step requires fitting independent logistic regression models for every genome-wide significant variant against ∼1000 clinical phenotypes across multiple cross-validation folds. To handle this heavily branched task space, we implemented a CPU-based distributed computing architecture. Utilizing the `joblib` library on multi-core compute nodes, we distributed the logistic regression workloads via symmetric multiprocessing. This allowed us to fully saturate available CPU resources, maximizing throughput when mapping the discovered genetic axes to specific clinical outcomes.

### UK Biobank Bulk Datasets

We utilized data from the UK Biobank (UKB), a large-scale prospective population-based cohort. Approximately 500,000 participants were recruited from across the United Kingdom between 2006 and 2010, with enrollment and data collection continuing through follow-up assessments. The analysis presented here was performed on all available datasets released up to December 2024.

We analyzed the following multi-modal data sources:

- **Abdominal MRI:** We utilized abdominal MRI scans from the first imaging visit (Instance 2), specifically the Dixon sequences (Field ID 20201) which provide 3D volumes with separated fat and water channels. The raw scans cover a field of view from the neck to the knee with dimensions of Depth=370, Height=224, Width=174 voxels. Following the protocol established by [2], we used 3D U-Net deep learning models trained to generate segmentation masks for nine distinct anatomical structures: lungs, spleen, pancreas, liver, heart, aorta, bladder, left kidney, and right kidney. Using these masks, each organ was extracted as a separate 3D image dataset. While the original scan volume is fixed, the extracted organ volumes vary in size based on the participant’s anatomy. This dataset included 81,321 participants for each abdominal organ.
- **Brain MRI:** For neuroimaging, we analyzed T1-weighted structural images of the brain (Field ID 20252). We utilized the processed NIfTI files provided by the UK Biobank, which have been subjected to a standardized processing pipeline including defacing, re-orientation, and cropping [75]. For this analysis, we used images that had been linearly and non-linearly registered to the standard MNI152 space, resulting in uniform 3D volumes with dimensions of (182, 182, 218). This dataset contained 59,070 participants.
- **Cardiac MRI (CMR):** We analyzed 3D cine MRI videos to capture dynamic cardiac function. We utilized two primary views: the long-axis (Field ID 20208) and short-axis (Field ID 20209) cine stacks. These sequences measure the variation of the heart chambers along and perpendicular to the long axis of the left ventricle, respectively, over the cardiac cycle. The final dataset included 79,898 participants with valid long-axis scans and 87,448 participants with valid short-axis scans. The video shapes were normalized to 16 x 48 x 128 x 128.
- **DXA Lateral Spine:** We utilized Dual-energy X-ray Absorptiometry (DXA) images of the lateral spine (Field ID 20158), which capture the lumbar and thoracic vertebrae (L4–T4). Quality control was performed to exclude scans exhibiting significant block artifacts or acquisition errors. These data consist of 2D images of varying shapes and, unlike the abdominal MRI data, were analyzed as whole images without prior segmentation. The final cohort included 69,418 participants.
- **Electrocardiogram (ECG):** We analyzed raw signal traces from the 12-lead resting electrocardiogram (ECG) recorded during the initial assessment center visit (Field ID 20205). The data consists of 12 voltage time-series (leads I, II, III, aVR, aVL, aVF, V1–V6) recorded at 500 Hz for a duration of 10 seconds.
- **Retinal Optical Coherence Tomography (OCT):** We utilized spectral-domain OCT volumetric scans of the left eye (Field ID 21017). Each scan consists of 128 cross-sectional B-scan images (512 × 650 pixels, grayscale), which together form a 3D volume capturing the layered microstructure of the retina, including the macula and surrounding regions. These volumetric scans enable the assessment of retinal layer thicknesses and structural abnormalities relevant to conditions such as glaucoma and macular degeneration. The final dataset included 66,915 participants.
- **Retinal Fundus Photography:** We analyzed color fundus photographs of both the left (Field ID 21015) and right (Field ID 21016) eyes. Quality-controlled images were resized to 384× 384 pixels (RGB). These 2D images capture the posterior pole of the eye, providing visualization of the optic disc, macula, and retinal vasculature. The final dataset included 80,658 participants.
- **Plasma Proteomics:** We utilized the Normalized Protein Expression (NPX) data from the UK Biobank Pharma Proteomics Project (UKB-PPP), (Field ID 30900). This panel measures the abundance of 2,922 circulating proteins using the Olink Explore 3072 Proximity Extension Assay (PEA). The data were processed according to the project’s white paper guidelines [16] without further modification. This dataset included 48,581 participants.
- **NMR Metabolomics:** We analyzed the nuclear magnetic resonance (NMR) metabolomics panel (Field ID 220), which quantifies 251 metabolic biomarkers including lipids, fatty acids, and low-molecular-weight metabolites. Detailed processing information is available in the UK Biobank documentation [6]. Our analysis dataset included 248,183 participants.

### Feature Extraction via Self-Supervised Learning

To generate representations (embeddings) of the diverse data modalities, we trained self-supervised deep learning models. Specifically, we leveraged Masked Image Modeling (MIM) as a primary strategy for the imaging modalities. MIM demonstrates strong capability in extracting generalizable semantic features from medical imaging data, scaling effectively from static 2D images to dynamic 3D videos [8, 76] while employing the same broad objective and model architectures.

#### Static Imaging (Abdominal MRI, Brain MRI, DXA, Retinal Imaging)

For anatomical imaging, we utilized a Masked Autoencoder (MAE) training objective to learn representations using a Vision Transformer Small (ViT) encoder. The final embedding dimension for all static imaging modalities was 384.

- **Abdominal and Brain MRI:** We adopted a slice-based encoding strategy (detailed in [77]). We utilized a modified ViT-b architecture (*d* = 384), where the patch size was adaptively optimized for each organ based on its average physical dimensions (see Supplement S8). The model was trained on 2D slices. At inference, embeddings were extracted directly from the encoder’s `[CLS]` token for each 2D axial slice (*H ×W*). To obtain a single representation for the entire 3D volume, we performed global average pooling across the depth (*z*-axis) dimension.
- **DXA:** We applied the same MAE training strategy to the 2D lateral spine scans. Since these are single 2D images, the embedding was derived directly from the encoder output without depth-wise pooling.
- **Retinal OCT:** We applied the same slice-based MAE strategy to the OCT B-scan volumes. Individual 2D B-scans were encoded independently, and a single 384-dimensional volume-level embedding was obtained via global average pooling across all 128 slices.
- **Retinal Fundus:** We trained an MAE on the 2D color fundus photographs (3-channel RGB, 384 ×384). The embedding was derived directly from the encoder output without depth-wise pooling.

#### Dynamic Imaging (Cardiac MRI)

To capture the spatiotemporal dynamics of cardiac function, we employed a VideoMAE framework [76] applied to both 3D short-axis and 2D long-axis cardiac videos [78]. The model processes 2D subvolumes (*H ×W ×T*) extracted from the cine cardiac MRI (cMRI) sequences, followed by global average pooling across *z*-axis for short-axis 3D cMRI. Unlike in static images, this model learns joint space-time features. At inference, we utilized global average pooling across the spatiotemporal patch embeddings to generate a single 768-dimensional feature vector per scan.

#### Physiological Time-Series (ECG)

For the 12-lead ECG data, we utilized a self-supervised model optimized for cardiac signal representation [79]. The model processes the raw voltage traces to extract a compact 768-dimensional embedding that captures morphological waveform characteristics independent of standard clinical intervals.

#### Molecular Assays

For the high-dimensional molecular assays, the normalized abundance measurements were treated as direct embedding equivalents, preserving the interpretability of the individual markers. We utilized the full vector of 2,922 normalized protein expression levels from Plasma Proteomics (PPP) and the vector of 251 metabolic biomarkers from NMR Metabolomics assays.

### Genetic Ancestry and Cohort Filtering

For all association analyses, we restricted the cohort to participants with “European” genetic ancestry (labeled as EUR in the Pan-UKBB nomenclature). Ancestry assignment was based on genetic principal component projections provided by the Pan-UK Biobank (Pan-UKBB) consortium [80]. We excluded any participants identified with sex chromosome aneuploidy.

For the NMR metabolomics analysis specifically, we applied an additional exclusion criterion: participants reporting the use of any statin medication were removed from the cohort to mitigate confounding effects. Medication usage was determined based on self-reports from `UK Biobank Field ID 20003` (Treatment/medication code).

### Genotype Data and Variant Processing

We utilized the UK Biobank version 3 imputed genotype dataset, which provides coverage of over 90 million variants imputed using the Haplotype Reference Consortium (HRC) and UK10K haplotype resources [6]. We lifted the variant calls to the GRCh38 assembly. Where available, we report the reference SNP rsID for known variants.

#### Variant Filtering and Selection

To ensure robust estimation of the covariance matrices required for our linear discriminant analysis (LDA) classifier, we filtered variants based on minor allele frequency (MAF). Specifically, we included all known coding variants with a minimum of 10 participants possessing the homozygous alternate genotype (*N*_hom-alt_ *≥*10) within our analytical cohort. This filtering strategy resulted in a final set of **917**,**921** variants for association testing in the imaging modalities. The precise number of variants included varied slightly for each data modality depending on the specific sample size of the respective cohort.

#### Computational Optimization

To optimize data read throughput and memory utilization during the high-dimensional association study, we performed a conversion from the standard BGEN format. Genotypes were extracted and serialized into Apache Parquet files using the PyArrow engine. Genotypes were encoded as integer values (0 for homozygous reference, 1 for heterozygous, 2 for homozygous alternate) and stored using int8 precision. This serialization strategy significantly reduced the I/O overhead associated with standard BGEN decompression and parsing.

#### Variant Annotation and Prioritization

To assign a functional context to each variant, we employed the Open Targets Variant-to-Gene (V2G) pipeline [81]. This disease-agnostic model generates a score for variant-gene pairs; we assigned each variant to the gene with the highest V2G score. Variant consequences were annotated using the Ensembl Variant Effect Predictor (VEP) version 100 [82]. Putative loss-of-function (pLoF) variants were identified using the LOFTEE plugin [83].

#### Linkage Disequilibrium Clumping

To identify independent biological signals among our associations, we grouped variants into independent loci based on Linkage Disequilibrium (LD). Following a greedy approach similar to the PLINK `clump` algorithm, we performed the following steps, run independently for each modality:

1. **Selection of Lead Variants:** We identify the most significant variant (*P <* 5× 10^*−*8^) across the genome to serve as a potential lead variant.
2. **LD-Based Aggregation:** We define an LD block by tagging all neighboring variants within a window of *±*500 kb that exhibit an LD *r*^2^ *>* 0.5 with the lead variant.
3. **Iterative Refinement:** These tagged variants are assigned to the lead variant’s locus and removed from the candidate pool. This process is repeated until no variants remains above the genome-wide significance threshold.

To ensure the LD structure accurately reflects our study population, we calculated *r*^2^ values using the genotype calls from the UK Biobank, restricted to the same subset of individuals of European genetic ancestry used in the primary association analysis. These LD-clumped results are used when reporting overall number of loci discovered and reporting top disease associations.

## Downstream Analysis

### Counting Significant Loci

After LD-clumping and applying a genome-wide significance threshold of *P <* 5 ×10^*−*8^, we count the number of independent significant loci within each modality. We consider two different models of variant effect (homozygous, dominant) as previously described, as well as a “combined” version of results where we pick the contrast with a lower p-value for each variant.

We also aggregate the significant loci across modalities using the following groups:

1. **All Organs**: 17 modalities - ECG, long-axis cardiac video, short-axis cardiac video, static heart imaging, aorta, lungs, liver, pancreas, spleen, left kidney, right kidney, bladder, spine DXA, brain T1, OCT left eye, OCT right eye, retinal fundus
2. **Primary Organs**: 9 modalities (one representative per major organ system) - short-axis cardiac video, lungs, liver, pancreas, spleen, right kidney, spine DXA, brain T1, OCT left eye
3. **Omics**: 2 modalities - PPP proteomics, NMR metabolomics

### Comparison to GWAS

To directly compare our results to GWAS, we defined four categories of quantitative traits derived from the same data sources as our primary cardiac, brain, protein and metabolite results:

1. **Cardiac Imaging Traits (short axis)**: 61 traits derived from short-axis cardiac MRI [41]. Accessed from UKB category 157, filtering to ventricular traits (“LV” or “RV” in the name) as other traits (atrial, aortic) are not measured from short-axis scans.
2. **Brain Imaging Traits (T1-weighted)**: 1436 traits derived from T1-weighted brain MRI [11]. Accessed from UKB category 110.
3. **Protein Abundances (PPP)**: 2913 traits from protein abundance [16]. Accessed from UKB field 30900.
4. **Metabolite Abundances (NMR)**: 251 traits from metabolite abundance [17]. Accessed from UKB field 220.

For each set of phenotypes, we subsequently ran *regenie* [84], using the same covariates and individuals that were used when running our V2V method on each of these four modalities. For step 1, we select high-quality SNPs with minor allele frequency (MAF) *<* 1%, minor allele count (MAC) *>* 100, genotype missingess *<* 10% and Hardy-Weinberg equilibrium *p <* 10^*−*15^. For step 2, we use the same variants that were used in our V2V method. We use an additive model and apply Rank-Inverse Normal Transform to the phenotypes in both steps. For comparison to V2V, we aggregated summary statistics within each phenotype group, picking the best Bonferroni-corrected p-value across traits for a given variant.

### Identifying Novel Associations

To identify novel associations, we compared our results to previously reported associations from GWAS Catalog [15] (downloaded on 2025-12-22). For each organ system, we define a set of relevant trait categories from the Experimental Factor Ontology [46], listed in Table 2.

**Table 2.** Organ-specific clinical traits and biomarkers.

| Organ | List of Traits / Categories |
| --- | --- |
| Bladder | bladder disease |
| Bone | bone density, bone disease, bone measurement |
| Brain | brain disease, neuroimaging measurement |
| Cardiac | cardiovascular disease, cardiovascular disease biomarker measurement, magnetic resonance imaging of the heart |
| Eye | eye disease, eye measurement |
| Kidney | cystatin C measurement, glomerular filtration rate, kidney disease, kidney volume |
| Liver | liver disease, liver disease biomarker, liver volume |
| Lungs | lung capacity, respiratory disease biomarker, respiratory system disease |
| Pancreas | pancreas disease, pancreas volume, pancreatic fat pad amount |
| Spleen | spleen iron amount, spleen volume, splenic disease |

For each organ system (excluding blood / omics), we identify genome-wide significant associations in GWAS catalog where the reported trait matches one of these traits (or any of its child traits defined in EFO). To account for linkage disequilibrium, we mark regions of *±*500 kilobases around any reported variant as a “previously associated region”. Then, we search our organ-specific results for significant variants that do not fall into any previously associated region.

## Data Availability

The UK Biobank data used in this study are available via the UKB research application process. Derived summary statistics will be made available at the time of peer-reviewed publication.

## Acknowledgements

This work was conducted using UK Biobank application number 44584.

