## Supplementary Material for "Vector2Variant: Discovery of Genetic Associations from ML Derived Representations without Phenotype Engineering"

#### Contents

**S1 Out-of-sample evaluation ensures calibrated statistics in high-dimensional multivariate analysis** **1**

**S2 V2V can detect rare variants from ClinVar below 1% MAF** **4**

**S3 Comparison to Existing Multivariate Association Methods** **6**

**S4 Embeddings capture subtle features and they may not align with the principal components** **9**

**S5 Summary of Novel Loci** **11**

**S6 Novel Association Examples** **11**

**S7 Abdominal Organ Segmentation from Dixon MRI** **17**

**S8 Masked AutoEncoder Model for Static MRI and DXA, Retinal Imaging** **18**

**S9 VideoMAE Model for Cardiac MRI** **21**

**S10 ECG Self-Supervised Representation Model (ST-MEM)** **22**

### Method Validation & Comparisons

#### S1 Out-of-sample evaluation ensures calibrated statistics in high-dimensional multivariate analysis

A critical challenge in multivariate genotype-phenotype analysis arises when the number of phenotypic features approaches or exceeds the number of samples in a genotype class. With hundreds to thousands of features (e.g., 500 for organ imaging embeddings, 3,000 for plasma proteomics) and often fewer than 50 carriers for rare variants, the ratio of dimensions to carriers can exceed 10:1, creating severe overfitting risk.

To demonstrate this, we tested 100,000 random variants stratified by carrier frequency for association with liver MRI embeddings and compared the resulting p-value distribution against the uniform null expectation using QQ plots. For each variant, we fit Linear Discriminant Analysis on a training set (40% of samples) and evaluated the learned projection on both the training set (in-sample) and a held-out validation set (out-of-sample). We quantified statistical calibration using the genomic control factor $\lambda_{\mathrm{GC}}$ [1]. On real data, in-sample (training) p-values were massively inflated ($\lambda_{\mathrm{GC}} = 1{,}375$), whereas out-of-sample (validation) p-values were near the expectation under the null ($\lambda_{\mathrm{GC}} = 1.15$) (Fig. S1, top row).

To confirm that the in-sample inflation reflects overfitting rather than true signal, we performed a permutation analysis. We sampled 1,000 variants stratified by carrier frequency and, for each variant, shuffled genotype labels 100 times to destroy any true genotype-phenotype association, yielding 100,000 permuted null tests. Under this null, in-sample testing produced extreme genomic control values ($\lambda_{\mathrm{GC}} = 144$), demonstrating that LDA can separate random groups when tested on training data (Fig. S1, bottom left). In contrast, out-of-sample p-values followed the expected uniform distribution ($\lambda_{\mathrm{GC}} = 1.001$), indicating proper Type I error control (Fig. S1, bottom right). The near-unity genomic control under permutation establishes a critical baseline: any departure from $\lambda_{\mathrm{GC}} \approx 1$ observed on real data in the out-of-sample setting can be attributed to genuine associations rather than statistical miscalibration.

Existing multivariate methods such as MultiPhen [2] and MOSTest [3] evaluate associations on the same data used for model fitting, relying on asymptotic properties of likelihood ratio tests to maintain calibration. These theoretical guarantees, however, require assumptions about the relationship between sample size and feature dimensionality that are difficult to verify and may not hold in modern high-dimensional settings. By explicitly partitioning data into training and validation sets, our approach sidesteps these assumptions entirely. The out-of-sample evaluation provides a direct, assumption-free test of whether learned multivariate signatures generalize beyond the data used to discover them.

Having established that out-of-sample evaluation eliminates spurious statistical artifacts, we verified that the framework produces well-calibrated test statistics across all imaging modalities. We computed out-of-sample $\lambda_{\mathrm{GC}}$ for all 17 organ modalities (Table S1; representative QQ plots in Fig. S2). Across all organs, validation $\lambda_{\mathrm{GC}}$ values ranged from 1.005 (OCT, left eye) to 1.652 (brain T1), with a median of 1.15. We repeated the permutation null analysis (1,000 variants $\times$ 100 permutations) independently for each organ; the resulting $\lambda_{\mathrm{GC}}$ values were tightly calibrated across all modalities (median = 1.0005, range: 0.999–1.001), indicating proper Type I error control regardless of organ system or sample size.

The variation in $\lambda_{\mathrm{GC}}$ across modalities is consistent with differences in the richness of genetic signal captured by each representation. Brain T1 ($\lambda_{\mathrm{GC}} = 1.65$) and short-axis cardiac video ($\lambda_{\mathrm{GC}} = 1.42$) are among the highest, reflecting the extensive genetic architecture of brain structure and cardiac function. Conversely, retinal OCT ($\lambda_{\mathrm{GC}} =$

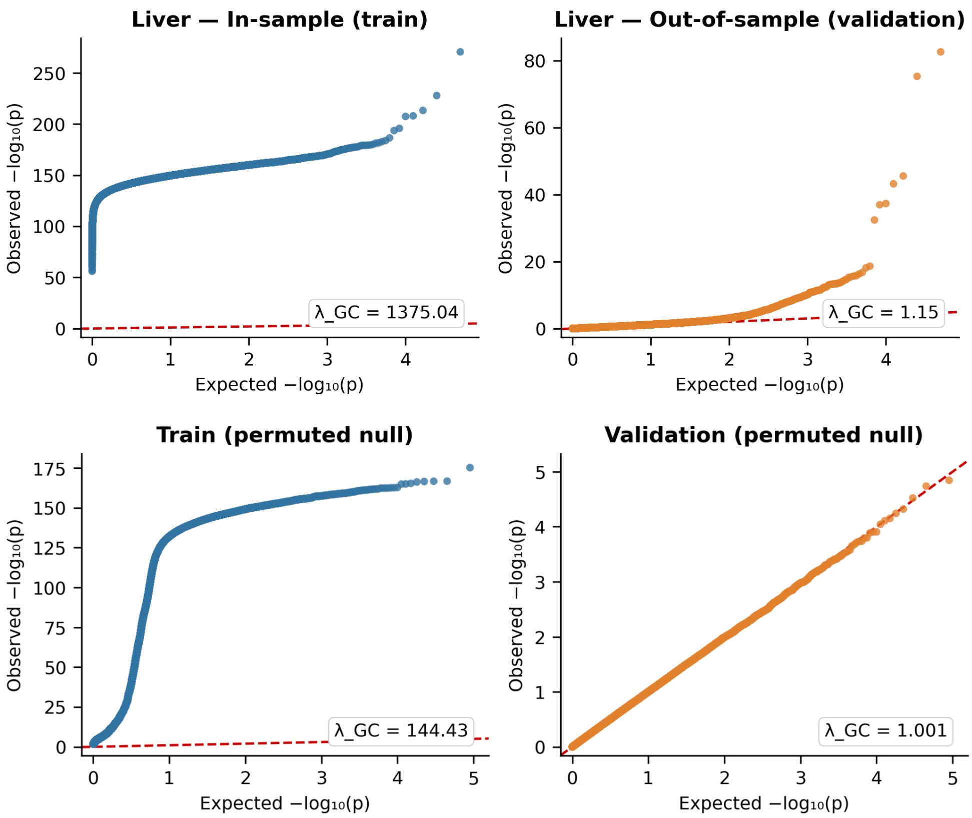


**Fig. S1 Train-validation split prevents statistical artifacts in high-dimensional settings.** QQ plots of $-\log_{10}(p)$ values for 100,000 random variants tested against liver MRI embeddings. The red dashed line indicates the expected uniform distribution under the null. **(Top row)** Real data. In-sample (train, left) evaluation shows massive inflation ($\lambda_{GC}$ = 1,375), with observed p-values departing from the null by over two orders of magnitude. Out-of-sample (validation, right) evaluation yields near-calibrated statistics ($\lambda_{GC}$ = 1.15), with modest departure from the diagonal reflecting genuine polygenic signal. **(Bottom row)** Permuted null (1,000 variants × 100 shuffled permutations each). In-sample evaluation (left) produces equally extreme inflation ($\lambda_{GC}$ = 144), confirming that LDA can separate random groups when evaluated on training data. Out-of-sample evaluation (right) yields properly calibrated p-values tightly following the diagonal ($\lambda_{GC}$ = 1.001), confirming correct Type I error control.

1.01–1.02) and pancreas ($\lambda_{GC}$ = 1.07) show lower values, consistent with fewer or sparser genetic associations for these modalities. Notably, all train $\lambda_{GC}$ values were uniformly extreme (range: 452–1,542) regardless of whether the modality harbored strong genetic signal, underscoring that in-sample evaluation provides no meaningful discrimination between true and spurious associations.

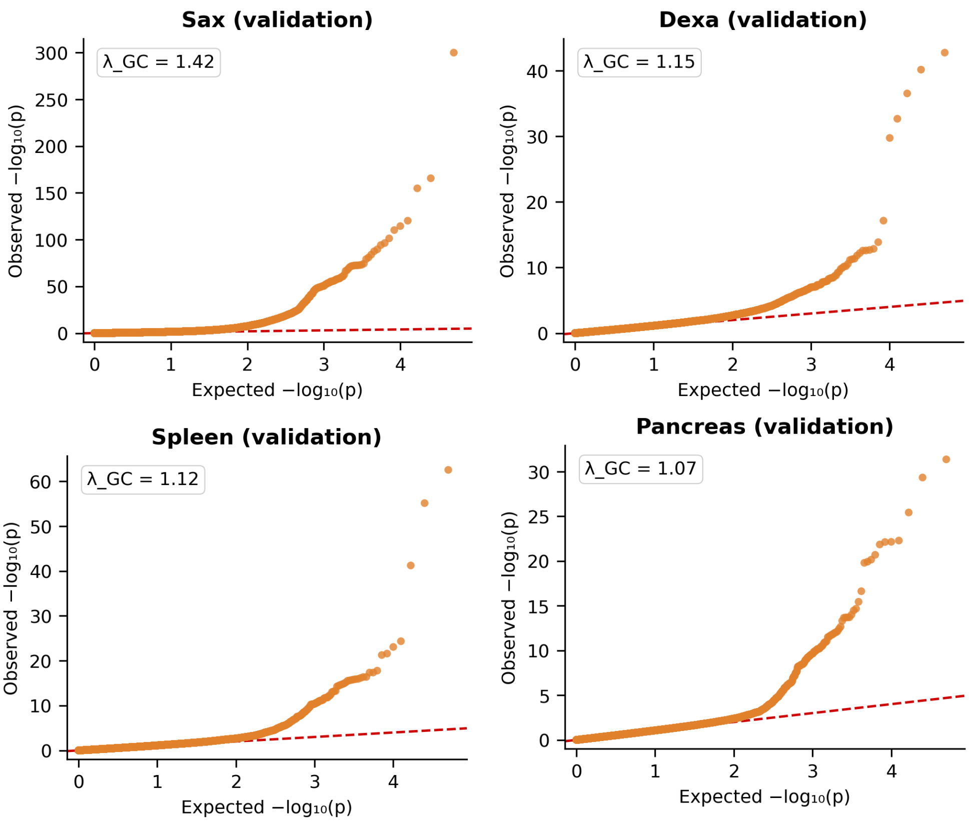


**Fig. S2 Out-of-sample QQ plots across representative organ systems confirm well-calibrated statistics.** QQ plots of $-\log_{10}(p)$ values from out-of-sample (validation) evaluation of 100,000 random variants for four representative modalities: short-axis cardiac video (SAX, $\lambda_{GC} = 1.42$), lateral spine DXA ($\lambda_{GC} = 1.15$), spleen ($\lambda_{GC} = 1.12$), and pancreas ($\lambda_{GC} = 1.07$). The red dashed line indicates the expected uniform null distribution.

### S2 V2V can detect rare variants from ClinVar below 1% MAF

In V2V, statistical significance is based on regressing projection score against allelic dosage (see Methods). The power to detect separation in the latent space is characterized by the same parameters as GWAS, mainly minor allele frequency and cohort size. By calculating continuous projection scores for the entire cohort, V2V leverages information from non-carrier individuals who "phenocopy" the genetic effect, which could potentially improve power to detect rare variants.

Empirically, the V2V framework demonstrates sensitivity for detecting rare variants, including those with a minor allele frequency (MAF) below 1%. In our Results, we describe our ability to capture known associations with low-frequency variants as the *PCSK9* rs11591147 ($MAF \approx 1.7\%$) from proteomics and *IL11* rs4252548

| **Modality** | $\lambda_{GC}$ **(validation)** |
|---|---|
| Brain (T1w) | 1.652 |
| Heart Video (SAX) | 1.419 |
| Heart | 1.270 |
| Heart Video (LAX) | 1.265 |
| Lungs | 1.257 |
| Aorta | 1.177 |
| ECG | 1.153 |
| DXA Lat. Spine | 1.152 |
| Left Kidney | 1.151 |
| Liver | 1.151 |
| Right Kidney | 1.147 |
| Spleen | 1.122 |
| Bladder | 1.091 |
| Pancreas | 1.065 |
| OCT (right eye) | 1.024 |
| Retinal Fundus | 1.019 |
| OCT (left eye) | 1.005 |

**Table S1** Out-of-sample genomic control factors ($\lambda_{GC}$) across all 17 imaging modalities, sorted by descending $\lambda_{GC}$. Values were computed from 100,000 random variants evaluated on held-out validation data. Permutation null analysis (1,000 variants × 100 permutations per organ) yielded tightly calibrated $\lambda_{GC}$ across all organs (median = 1.0005, range: 0.999–1.001), indicating proper calibration under the null hypothesis.

($MAF \approx 2.2\%$) from spine DXA. Here, we highlight our detection of several high-impact, low-MAF coding risk alleles documented in the ClinVar database [4]:

- ***TNFRSF13B*** **rs34557412** (MAF ≈ 0.7%): The projection axis optimized for this missense variant was highly significant within spleen embeddings ($-\log_{10} P \approx 17$). Our ProjWAS highlighted hematologic conditions and blood cancers (Fig. S3a). Notably, the top disease by effect size was common variable immunodeficiency (CVID) ($-\log_{10} P \approx 4.2$ , $\beta \approx 3.4$) an established ClinVar risk association for this allele [5], which V2V successfully recovered despite there being only $N = 5$ diagnosed cases in our cohort .
- ***CSRP3*** **rs45550635** (MAF ≈ 0.5%): We observed highly significant genotype separation within short-axis (SAX) cardiac video embeddings ($-\log_{10} P \approx 36$) for this missense variant. While ClinVar classifies this as a known cardiomyopathy risk allele [6], our ProjWAS of the resulting projection score revealed robust clinical associations with atrial fibrillation, hypertension, and chronic ischemic heart disease (Fig. S3b).

- ***SERPINA1* rs28929474** (MAF $\approx$ 2.0%): This variant , a known risk allele for Alpha-1 Antitrypsin Deficiency (AATD) [7], exhibited significant genotype separation in liver embeddings ($-\log_{10} P \approx 45$) and in lung embeddings ($-\log_{10} P \approx 7.8$). The resulting projection phenotypes yielded clinical associations consistent with AATD pathology [8], including fibrosis and cirrhosis of the liver, type 2 diabetes, and chronic obstructive pulmonary disease (COPD), with the latter association replicated across both liver- and lung-derived scores in our analysis (Fig. S3c).

These results suggest that V2V can surface pathogenic variation in high-dimensional data even for very rare variants, and subsequently identify relevant disease associations even when clinical diagnostic labels are extremely sparse.

### S3 Comparison to Existing Multivariate Association Methods

Several classes of methods have been developed to move beyond univariate GWAS by jointly analyzing multiple phenotypes. These approaches broadly fall into three categories: dimensionality reduction followed by GWAS, multivariate omnibus tests, and reverse regression models. While each addresses a limitation of standard univariate testing, they share common weaknesses that our framework is designed to overcome. First, all existing methods operate on a pre-defined set of scalar phenotypes; to our knowledge, our work is the first to demonstrate the ability to obtain genetic associations directly from neural network embeddings of raw high-dimensional data. Second, none learn a *variant-specific* axis of phenotypic variation—they either test a fixed set of components for every variant or produce an omnibus statistic without an interpretable direction. Third, methods that do fit models in the phenotype space rely on asymptotic calibration guarantees that degrade when the number of features approaches or exceeds the number of carriers, a regime that is common for rare variants in high-dimensional embeddings (Supplementary Section S1). Our method addresses all three limitations: it operates directly on embedding vectors without pre-defined phenotypes, learns a discriminative axis tailored to each variant, and uses an explicit train-validation split to ensure calibrated statistics regardless of the feature-to-sample ratio.

#### Dimensionality Reduction Approaches

A natural strategy for handling high-dimensional phenotypic data is to first reduce it to a small number of components and then perform univariate GWAS on each component. Principal component analysis (PCA) is the most common choice and has been applied at scale to UK Biobank imaging-derived phenotypes [9, 10] and to molecular panels [11]. Related approaches include independent component analysis (ICA) [12] and Bayesian factor models such as FactorGo [13].

The fundamental limitation of these approaches is that they identify axes of **maximum variance**, not axes of **genotypic separation**. In biobank-scale datasets, the dominant sources of variance in imaging embeddings or molecular panels are demographic confounders such as age, sex, and BMI. A genetic variant that induces a subtle

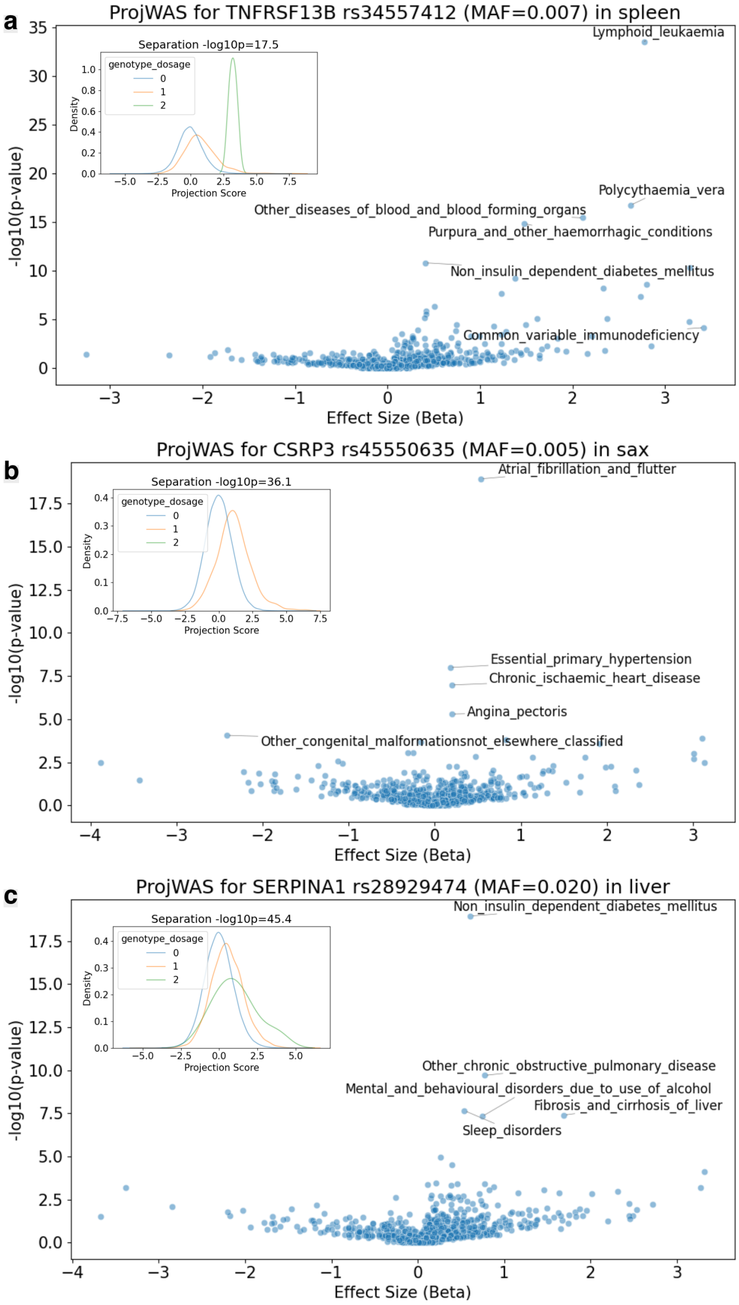


**Fig. S3 V2V recovers rare pathogenic variants documented in ClinVar.** **a**, ProjWAS volcano plot for the *TNFRSF13B* variant (rs34557412, $MAF \approx 0.7\%$) in spleen embeddings. The learned axis captures a known association with common variable immunodeficiency (CVID) as the highest effect size result, despite the extreme sparsity of diagnostic labels ($N = 5$ cases) in the cohort. **b**, ProjWAS volcano plot for the *CSRP3* missense variant (rs45550635, $MAF \approx 0.5\%$) derived from short-axis cardiac video embeddings. The projection phenotype significantly associates with cardiovascular diseases, consistent with its ClinVar classification as a cardiomyopathy risk allele. **c**, ProjWAS volcano plot for the *SERPINA1* variant (rs28929474, $MAF \approx 2.0\%$) in liver embeddings. The projection score recovers clinical signatures of Alpha-1 Antitrypsin Deficiency, including liver fibrosis, cirrhosis, and chronic obstructive pulmonary disease (COPD). In all subpanels, the inset panel at upper left shows a normalized kernel density plot of the V2V projection scores for individuals of each genotype dosage, showing that we can separate carriers from non-carriers despite the low allele frequency.

but biologically meaningful change in embedding space, particularly a rare variant affecting a small subpopulation, will project weakly onto the top principal components and may be undetectable. Moreover, the same fixed set of components is tested for every variant, precluding the discovery of variant-specific phenotypic signatures. Finally, the choice of how many components to retain is arbitrary and introduces a bias-variance tradeoff: too few components lose signal, while too many reintroduce the multiple testing burden.

In contrast, our method identifies a bespoke projection axis for each variant by directly maximizing genotypic separation via Linear Discriminant Analysis. This axis may lie in a low-variance subspace that PCA would discard, yet still capture a genuine biological perturbation.

#### Multivariate Omnibus Tests

Multivariate omnibus methods test whether *any* linear combination of phenotypes is associated with a variant, producing a single aggregate test statistic. Classical approaches include MANOVA [14] and canonical correlation analysis (CCA) [15]. More recent methods designed for GWAS at scale include MOSTest [3], which decorrelates phenotypes via a rotation matrix estimated from the data, performs univariate GWAS on each rotated phenotype, and combines the results into a chi-squared omnibus statistic calibrated under permutation. MOSTest has been applied extensively to brain imaging phenotypes and has demonstrated substantial gains in locus discovery over univariate testing.

These methods are effective at boosting discovery power, but they share two limitations. First, they are *omnibus* tests: they report whether a multivariate association exists, but do not identify the direction of the effect in phenotype space. This means they cannot produce a per-individual continuous score for downstream analysis such as ProjWAS, limiting biological interpretability. Second, classical multivariate tests (MANOVA, Hotelling's $T^2$) require the sample size to substantially exceed the number of features ($N \gg D$) for the test statistic to follow its assumed distribution. In settings where $D$ approaches the number of carriers, common when applying high-dimensional embeddings to rare variants, the empirical covariance matrix becomes ill-conditioned or singular, and the test statistic no longer follows its assumed asymptotic distribution. MOSTest mitigates this through permutation-based calibration, but still requires predefined phenotypes and does not produce interpretable per-variant axes.

Our method addresses the ill-conditioning problem through Ledoit-Wolf shrinkage regularization of the within-class covariance matrix, and produces both a calibrated association statistic and an interpretable projection axis for each variant, while the train-val split ensures proper calibration regardless of the $D$-to-$N$ ratio.

#### Reverse Regression

Reverse regression methods invert the standard GWAS model by regressing genotype on phenotypes rather than phenotype on genotype. The most established method in this category is MultiPhen [2], which fits a proportional odds ordinal regression model predicting the ordered genotype $(0, 1, 2)$ from a set of phenotypes. The joint significance of the phenotype predictors is assessed via a likelihood ratio test. This formulation is conceptually closest to our approach, as both methods ask whether phenotypic features can discriminate between genotype classes.

The critical difference lies in statistical calibration. MultiPhen evaluates the likelihood ratio statistic on the *same* data used to fit the model, relying on asymptotic $\chi^2$ approximations to control the Type I error rate. As we demonstrate in S1, this assumption fails catastrophically in high-dimensional settings: when the number of features approaches the number of minority-class samples, in-sample test statistics are inflated by over 100 orders of magnitude relative to properly calibrated out-of-sample statistics. Our method explicitly guards against this by learning the discriminative axis on a training partition and evaluating significance exclusively on held-out data, providing assumption-free calibration. Additionally, while MultiPhen can in principle operate on high-dimensional feature vectors, to our knowledge it has only been applied to pre-defined scalar phenotypes and its calibration properties in high-dimensional embedding spaces remain unvalidated. Furthermore, it does not produce continuous scores for downstream interpretation via ProjWAS.

#### S4 Embeddings capture subtle features and they may not align with the principal components

First, we demonstrate that subtle anatomical phenotypes, such as DISH scores, can be accurately regressed from lateral DXA image embeddings, even when the underlying representation learning models are not explicitly optimized to capture them. As illustrated in Fig. S4 (top), the Spearman rank correlation between ground-truth and predicted DISH scores on a held-out test set steadily improves as a function of training set size. This confirms that the latent embedding space encodes sufficient phenotypic information to identify a targeted projection axis highly correlated with the DISH score (reaching an $r = 0.52$).

Next, we investigate whether the principal components (PCs) of these lateral DXA embeddings naturally align with biologically relevant features. Using the genetic variant rs4252548 and its established association with lateral DXA-derived DISH scores [16] as an illustrative example, we find that standard unsupervised variance maximization is insufficient for genetic discovery. Specifically, the rank correlation between the DISH score and any of the top 50 PCs remains below 0.15, despite these components collectively explaining $> 90\%$ of the total variance in the embeddings. Furthermore, when we fit a linear model $p_{i,j} = \beta_v g_{i,v} + \epsilon_i$ (where $j \in \{1, \ldots, 50\}$ represents the PC index and $v$ represents the rs4252548 genotype), none of the top PCs reach genome-wide significance (Fig. S4, bottom). In contrast, because our V2V framework explicitly optimizes for a projection axis that maximizes allelic separation (Fig. **??**), it is uniquely equipped to uncover these subtle genetic associations, provided the requisite signal exists within the latent space. Once identified, these biologically aligned scalar projections can be utilized for downstream interpretation via ProjWAS.

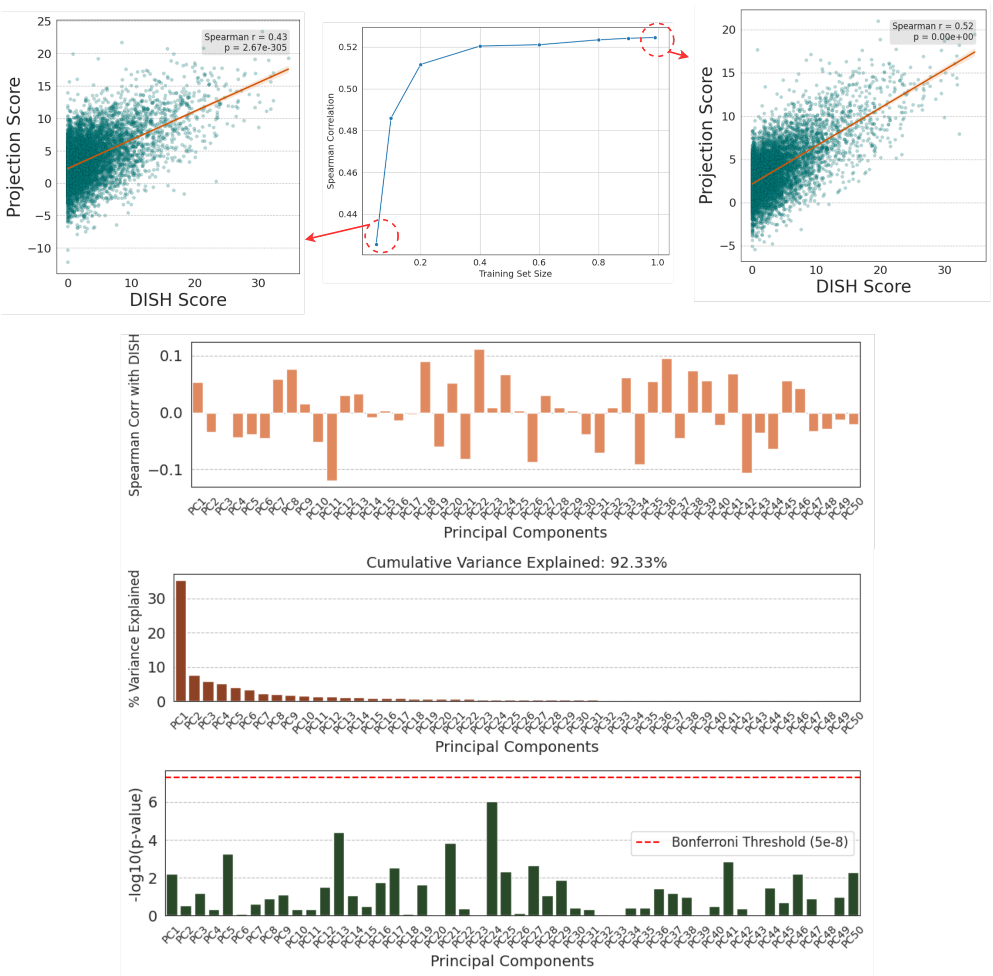


**Fig. S4 Targeted projections of DXA embeddings capture complex bone phenotypes better than unsupervised principal components.** (Top) Evaluation of linear regression models trained to predict DISH scores from lateral DXA embeddings. The central line plot illustrates the Spearman rank correlation on a held-out test set as a function of the training set size fraction. The flanking scatter plots detail the relationship between ground-truth DISH scores and predicted projection scores at low (left, $r = 0.43$) and high (right, $r = 0.52$) training data volumes. (Bottom) Unsupervised principal components (PCs) fail to isolate biologically relevant signals. The first bar plot shows the Spearman correlation between the top 50 PCs and the DISH score, demonstrating weak associations ($r < 0.15$). The second bar plot shows the percentage of variance explained by each PC, with the top 50 collectively accounting for 92.33% of the total variance. The third bar plot displays the $-\log_{10}(P\text{-value})$ for the association between the rs4252548 variant and each of the top 50 PCs; the red dashed line indicates the Bonferroni significance threshold. None of the top unsupervised PCs achieve genome-wide significance for this known variant-phenotype pair

### Novel Associations

#### S5 Summary of Novel Loci

As detailed in Methods, we identify novel associations which are not linked to relevant organ-specific traits in GWAS Catalog. We conducted a search for each of the 10 major organ systems studied, with results summarized in Table S2. To define the set of "system-level" associations, we combine significant results from both contrasts (homozygous, dominant) and group all modalities from the same organ system together (as defined in Table 1); if the same lead variant occurs multiple times within a given set, it is only counted once.

| Organ System | Novel Loci |
|---|---:|
| Eye | 131 |
| Brain | 83 |
| Spleen | 66 |
| Liver | 60 |
| Bladder | 36 |
| Kidney | 32 |
| Lungs | 29 |
| Cardiac | 27 |
| Pancreas | 8 |
| Bone | 2 |
| **Total** | **474** |

**Table S2** Number of novel primary loci identified in each organ system.

#### S6 Novel Association Examples

We highlight several additional examples of genes with at least one significant novel gene-organ association not found in GWAS Catalog. For each example, the linked figure includes LocusZoom plots and top-disease-ProjWAS results for each significant organ (**bold** indicates a novel organ association):

- **LRRC37A2 (S5)**: significant in **spleen**, brain, heart, kidney, lungs.
- **CATSPER4 / CNKSR1 (S6, S7)**: significant in **eye**, heart.
- **PCK2 (S8)**: significant in **kidney**, liver.
- **ITGA11 (S9)**: significant in **brain**.
- **SUPT3H / RUNX2 (S10)**: significant in **heart**, bone, brain.
- **UBE2U (S11)**: significant in **heart**.
- **USP37 (S12)**: significant in **brain**, **heart**.

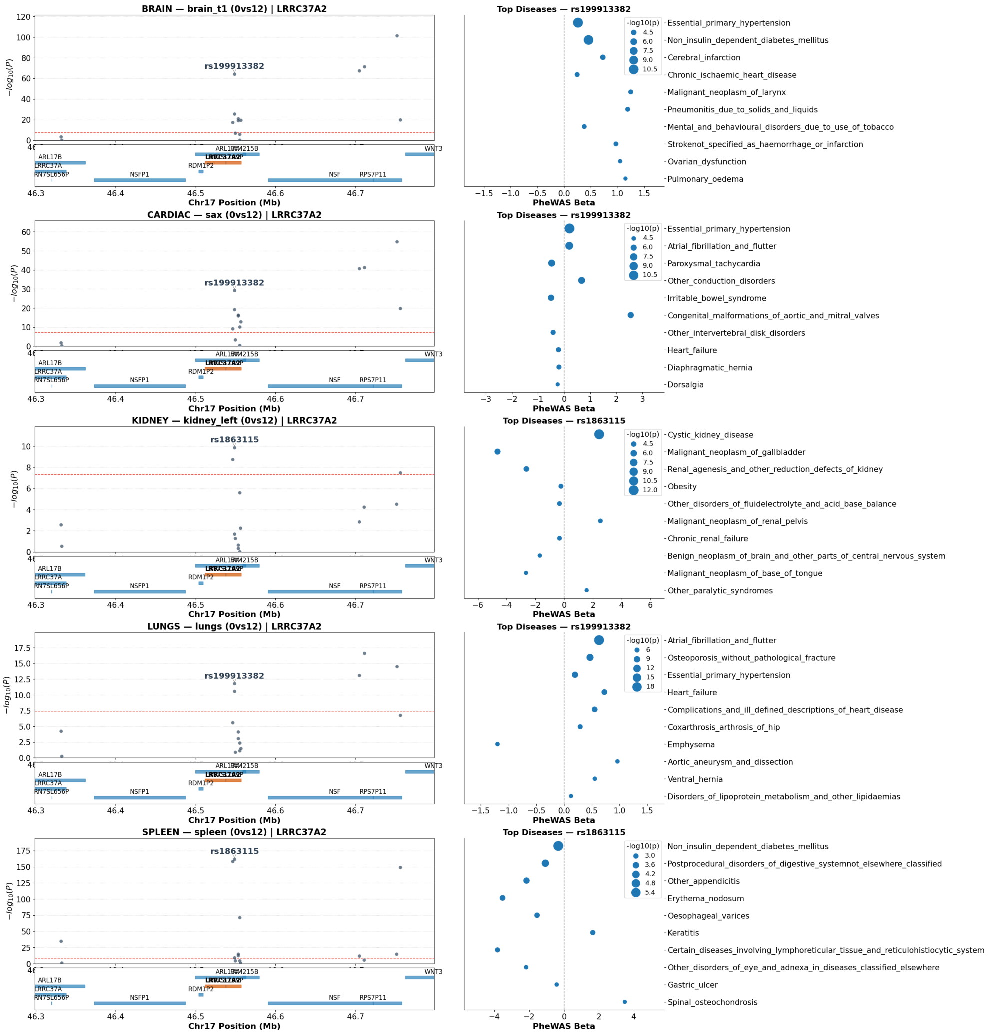


**Fig. S5 Top Organ-Variant-Disease Associations in LRRC37A2**

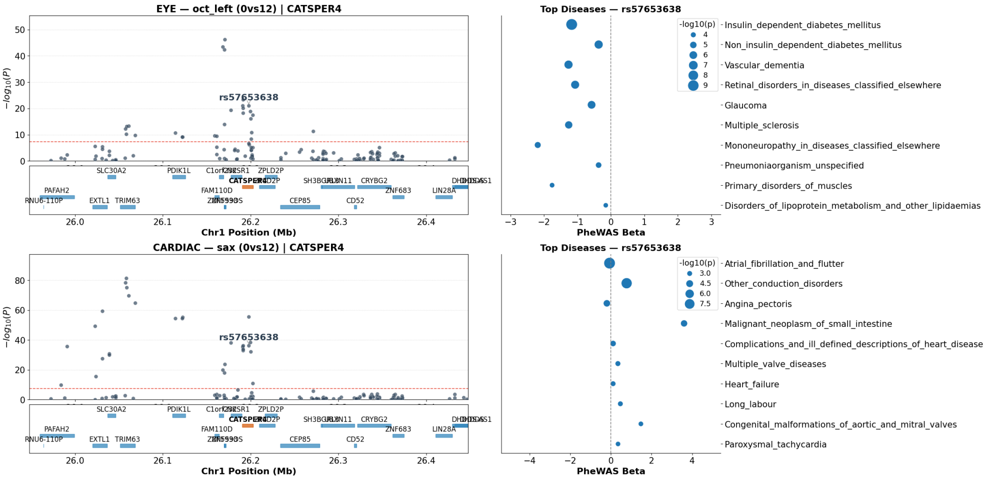


### Fig. S6 Top Organ-Variant-Disease Associations in CATSPER4

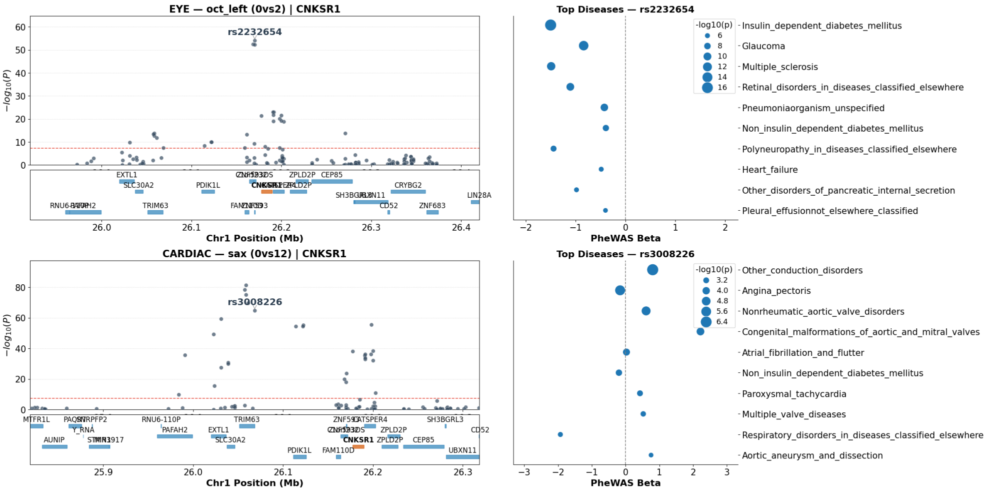


### Fig. S7 Top Organ-Variant-Disease Associations in CNKSR1

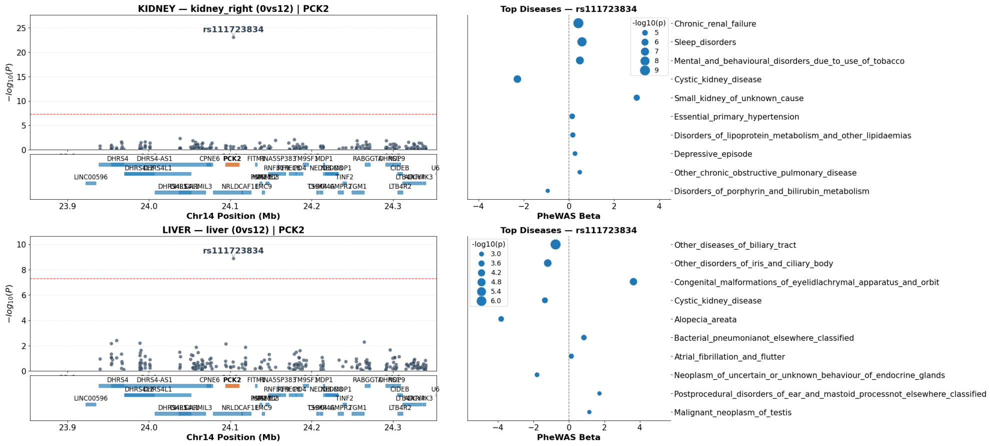


**Fig. S8** Top Organ-Variant-Disease Associations in PCK2

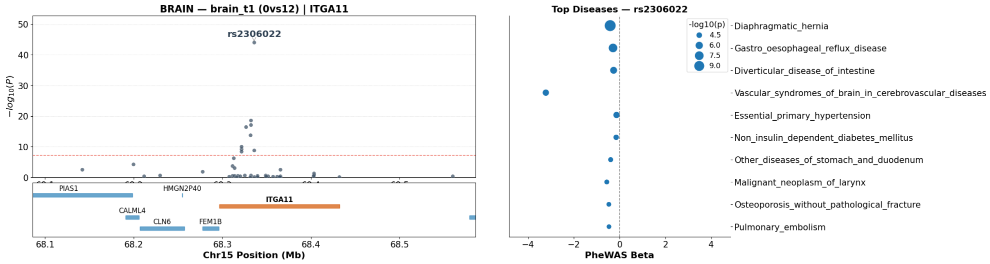


**Fig. S9** Top Organ-Variant-Disease Associations in ITGA11

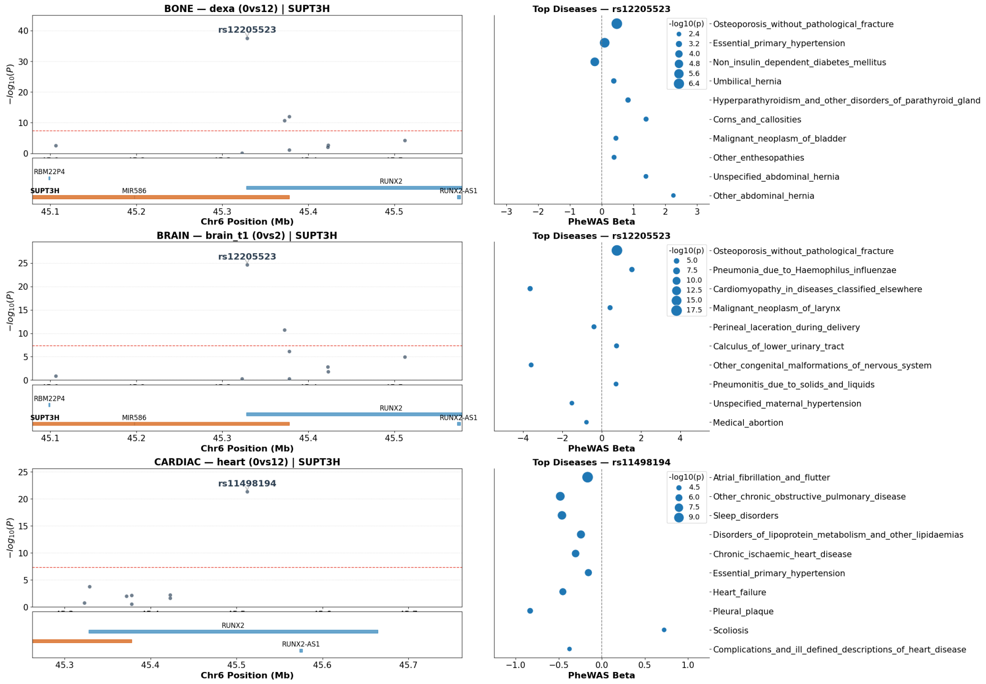


**Fig. S10 Top Organ-Variant-Disease Associations in SUPT3H / RUNX2**

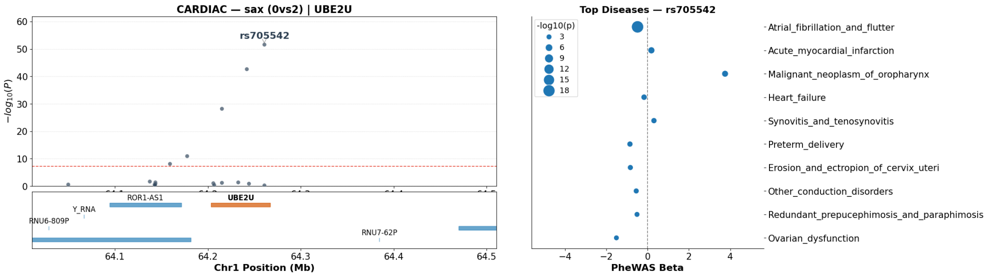


**Fig. S11 Top Organ-Variant-Disease Associations in UBE2U**

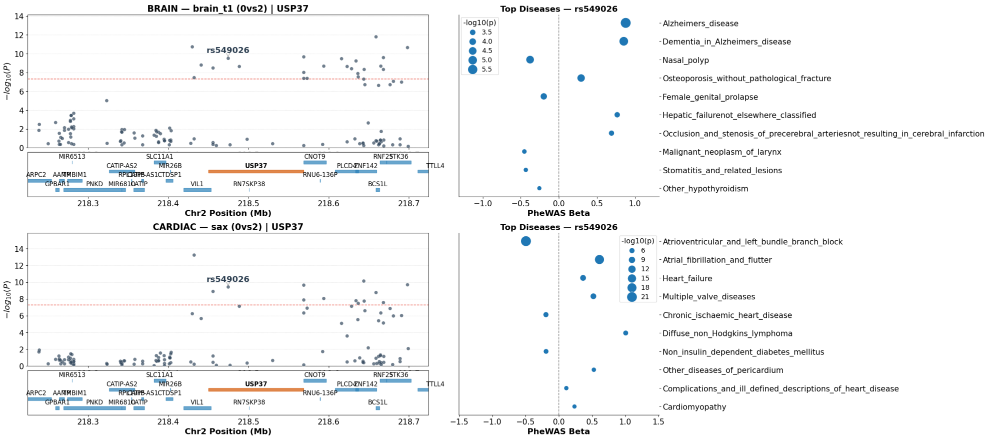


**Fig. S12 Top Organ-Variant-Disease Associations in USP37**

### Details of Segmentation & Embedding Models

#### S7 Abdominal Organ Segmentation from Dixon MRI

To isolate individual organs from the neck-to-knee abdominal Dixon MRI acquisitions (Field ID 20201), we developed a 3D deep learning segmentation pipeline that generates binary masks for nine anatomical structures: liver, heart, lungs, spleen, pancreas, aorta, bladder, left kidney, and right kidney. Each organ was then extracted as an independent 3D image dataset for downstream self-supervised representation learning.

##### Manual Annotation

Training labels were generated by manual annotation of abdominal structures by trained radiographers using MITK (mitk.org). Annotation channels were selected based on tissue contrast: water channels were used for organ delineation and fat channels for adipose-containing structures. Datasets for annotation were drawn from a sex-balanced cohort spanning a broad range of BMI, age, and ethnicity. Approximately 100-200 volumes were annotated per organ and used as ground truth. All annotations underwent quality control by a single expert with over 10 years of experience; annotations not meeting predefined criteria were rejected and re-annotated.

##### Architecture

We employed a 3D U-Net architecture with five encoding and four decoding blocks connected by skip connections [17]. Each encoding block comprised two $3 \times 3 \times 3$ convolutional layers with instance normalization and ReLU activation, followed by $2 \times 2 \times 2$ max-pooling. Feature channels doubled at each level from 32 to 512 at the bottleneck. Decoding blocks used transposed convolutions for upsampling, concatenated with the corresponding encoder skip connection, followed by two convolutional layers. The network accepted five input channels derived from the Dixon acquisition: fat, water, in-phase, out-of-phase, and body mask. Input volumes were zero-padded from the native $370 \times 174 \times 224$ voxels to $384 \times 176 \times 224$ to ensure divisibility by the pooling factors. Each model terminated in a binary classification head ($1 \times 1 \times 1$ convolution, two output channels).

##### Training Strategy

Training proceeded in two stages. First, a multi-task model with a shared encoder-decoder backbone and independent binary classification heads was trained to simultaneously segment all target organs, using a combined soft Dice and binary cross-entropy loss. This pre-training stage learned a general anatomical feature representation across structures. In the second stage, organ-specific models were initialized from the pre-trained backbone with randomly initialized classification heads and fine-tuned using soft Dice loss alone. This transfer learning approach enabled robust segmentation even for structures with limited annotations.

All models were optimized with Adam (learning rate $4 \times 10^{-3}$, exponential decay $\gamma$=0.99 per epoch) using mixed-precision (FP16) arithmetic. Elastic deformation was the primary data augmentation: a coarse random displacement field ($\sigma$=0.02) was sampled, upsampled via trilinear interpolation, and applied to both image and mask. No rotation or flipping augmentations were used, as the Dixon acquisition maintains consistent anatomical orientation. Training data were split 80/20 for training and validation. Pre-training ran for up to 150 epochs with early stopping; fine-tuning converged within 30 epochs. The final checkpoint for each organ was selected based on lowest validation loss, with ties broken in favor of earlier iterations.

##### Post-Processing and Quality Control

Predicted probability maps were thresholded at 0.5 to produce binary masks. Connected component analysis retained only the largest connected region for unpaired organs (e.g., liver, spleen, pancreas) and the two largest components for bilateral structures (kidneys, lungs). Quality control was performed by manual inspection of the 50 largest, 50 smallest, and 50 randomly sampled segmentations per organ. Cases with errors were documented, additional training annotations were generated, and models were retrained iteratively until visual QC criteria were satisfied.

##### Segmentation Performance

The ten organ-specific models achieved validation Dice scores ranging from 0.82 (pancreas) to 0.94 (heart), with a mean Dice score of 0.90 across structures (Fig. S13, left). Large, high-contrast structures such as the heart (0.94), lungs (0.93), liver (0.92), and spleen (0.92) achieved the highest scores. The pancreas, a small and irregularly shaped organ with low Dixon contrast, had the lowest score at 0.82 but still provided clinically useful segmentations. Bilateral kidney segmentation was consistent (left: 0.91, right: 0.90). Training and validation Dice scores showed close agreement across all structures (Fig. S13, right), indicating minimal overfitting.

#### S8 Masked AutoEncoder Model for Static MRI and DXA, Retinal Imaging

We employed a Masked Autoencoder (MAE) [18] framework to learn self-supervised representations from static imaging modalities: ten segmented abdominal organs from Dixon MRI, T1-weighted brain MRI, and DXA lateral spine images. Each modality was trained with an independent model instance.

##### Encoder Architecture

The encoder is based on a modified Vision Transformer Base (ViT-B) architecture. Starting from the standard ViT-B configuration ($d$=768, 12 layers, 12 attention heads), we applied two modifications to reduce computational cost and enable single-GPU training: (1) the embedding dimension was halved to $d$=384, and (2) the number

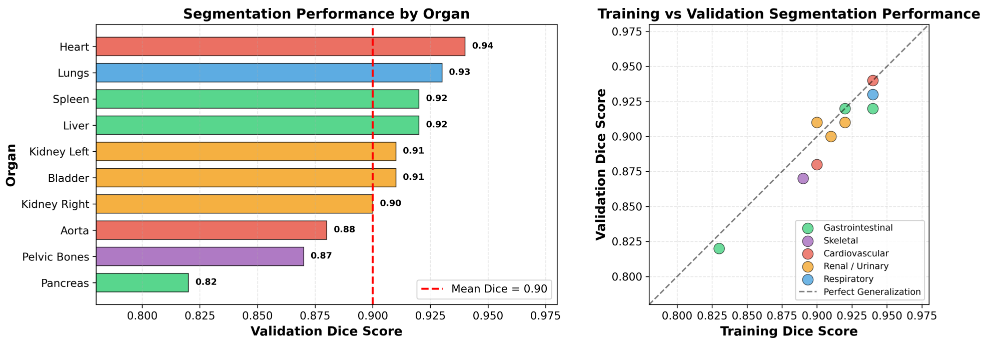


**Fig. S13 3D segmentation performance for the ten abdominal organs used in this study. (Left)** Validation Dice scores for each organ-specific U-Net model, colored by organ system. The dashed red line indicates the mean Dice score across structures (0.90). **(Right)** Training versus validation Dice scores for each organ. Points close to the diagonal (dashed line) indicate good generalization with minimal overfitting.

of transformer blocks was reduced from 12 to 8. The number of attention heads was kept at 12, resulting in a per-head dimension of 32 (compared to the standard 64). The MLP ratio remained at 4, giving a feed-forward hidden dimension of 1536. Fixed 2D sine-cosine positional embeddings [18] were used throughout.

#### Decoder Architecture

The decoder follows the asymmetric design of [18], with an embedding dimension of 512, 16 attention heads, and 6 transformer blocks. A linear projection maps encoder outputs ($d$=384) into the decoder space ($d$=512). Masked positions are filled with a shared learnable mask token, and both visible and masked tokens receive fixed 2D sine-cosine positional embeddings before decoding. The final prediction head is a linear layer projecting to $p^2 \times C$ values, where $p$ is the patch size and $C$ is the number of input channels.

#### Adaptive Patch and Image Sizing

Unlike standard MAE implementations that use a fixed $16 \times 16$ patch size on $224 \times 224$ images, we optimized both the image resize resolution and patch dimensions per organ to match the physical scale and spatial extent of each anatomical structure. Smaller organs (e.g., kidneys) were resized to smaller images with finer patches to preserve local detail, while larger structures (e.g. brain) used coarser patches at higher resolution. Table S3 summarizes the per-organ configuration.

#### Training Procedure

All models were optimized with AdamW [19] ($\beta_1$=0.9, $\beta_2$=0.95, weight decay =0.05). The learning rate followed a linear warmup schedule over 20 epochs, followed by half-cycle cosine decay to zero. The base learning rate was $1 \times 10^{-3}$, scaled linearly with effective batch size as $\text{lr} = \text{blr} \times \text{batch\_size}/256$. Abdominal organ models were trained

| Organ / Modality | Img Size | Patch | Grid | Chan. | Mask | Batch |
|---|---|---|---|---|---|---|
| Liver | 144 | 8 | 18 × 18 | 2 | 0.85 | 128 |
| Heart | 144 | 8 | 18 × 18 | 2 | 0.75 | 128 |
| Lungs | 180 | 12 | 15 × 15 | 2 | 0.85 | 128 |
| Spleen | 72 | 6 | 12 × 12 | 2 | 0.75 | 384 |
| Pancreas | 72 | 6 | 12 × 12 | 2 | 0.75 | 384 |
| Aorta | 72 | 6 | 12 × 12 | 2 | 0.85 | 512 |
| Bladder | 72 | 6 | 12 × 12 | 2 | 0.75 | 384 |
| Left Kidney | 48 | 6 | 8 × 8 | 2 | 0.75 | 1024 |
| Right Kidney | 48 | 6 | 8 × 8 | 2 | 0.75 | 1024 |
| Brain (T1w) | 192 | 8 | 24 × 24 | 1 | 0.75 | 64 |
| DXA Lat. Spine | 240 | 8 | 30 × 30 | 1 | 0.75 | 32 |
| OCT (B-scan) | 192 | 8 | 24 × 24 | 1 | 0.75 | 64 |
| Retinal Fundus | 384 | 16 | 24 × 24 | 3 | 0.75 | 32 |

**Table S3** Per-organ MAE configuration. Image size and patch size were adapted to each organ's spatial extent. Grid size denotes the number of patches per spatial dimension (img_size/patch_size). Abdominal organs use 2-channel input (Dixon water and fat); brain and DXA use single-channel input. All models share the same encoder ($d$=384, 8 blocks, 12 heads) and decoder ($d$=512, 6 blocks, 16 heads) architecture, with the exception of retinal fundus which uses a deeper encoder (12 blocks) and decoder (8 blocks). architecture.

for 100 epochs; the brain model was trained for 200 epochs to account for its smaller cohort and larger spatial extent. The DXA model was trained for 100 epochs with a warmup of 40 epochs and per-patch normalized pixel loss [18].

The reconstruction objective is the mean squared error (MSE) between predicted and ground-truth pixel values, computed only over masked patches. During training, a random subset of patches (determined by the mask ratio in Table S3) is removed from the input; the encoder processes only the visible patches, and the decoder reconstructs the full image. Weight initialization follows [18]: Xavier uniform for linear layers, and truncated normal ($\sigma$=0.02) for the `[CLS]` and mask tokens.

#### Slice-Based Encoding for 3D Volumes

For the 3D volumetric modalities (abdominal and brain MRI), we adopted a slice-based encoding strategy [20]. During both training and inference, 2D axial slices ($H \times W$) were extracted from the 3D volume and processed independently by the MAE. At inference, the `[CLS]` token embedding was extracted from each slice with no masking applied, yielding a 384-dimensional vector per slice. A single embedding for the entire 3D volume was obtained by global average pooling across the depth ($z$-axis) dimension. DXA and retinal fundus images, being inherently 2D, were encoded directly without depth-wise pooling. For OCT, the same slice-based strategy was applied: individual B-scan slices were encoded independently, and a single volume-level embedding was obtained by global average pooling across all 128 slices.

### S9 VideoMAE Model for Cardiac MRI

To model the spatiotemporal dynamics of cardiac function from cine MRI, we employed a VideoMAE [21] applied to both 3D short-axis and 2D long-axis cardiac videos [22]. The model processes 2D subvolumes ($H \times W \times T$) extracted from the cine cardiac MRI (cMRI) sequences, followed by global average pooling across $z$-axis for short-axis 3D cMRI. Unlike the slice-based MAE used for static imaging, VideoMAE processes spatiotemporal volumes jointly, enabling the model to learn representations that capture both anatomical structure and motion across the cardiac cycle.

#### Architecture

The encoder follows the standard ViT-B architecture: embedding dimension $d$=768, 12 transformer blocks, 12 attention heads, and MLP ratio of 4. The input video is tokenized via a 3D convolutional patch embedding with spatial patch size $8 \times 8$ and temporal tubelet size 2, yielding spatiotemporal tokens from an input of shape $3 \times 16 \times 128 \times 128$ (channels $\times$ frames $\times$ height $\times$ width). This produces $8 \times 16 \times 16 =$ 2,048 tokens per video. Fixed sinusoidal position embeddings encode both spatial and temporal location.

The decoder is lightweight relative to the encoder: embedding dimension 384, 6 attention heads, and 4 transformer blocks. A linear projection maps encoder outputs ($d$=768) into the decoder space ($d$=384), where learnable mask tokens are inserted at masked positions for reconstruction.

#### Masking Strategy

VideoMAE employs a two-stage masking scheme. During training, **tube masking** at 90% ratio removes entire spatiotemporal tubes (consistent across all temporal frames for a given spatial position), ensuring the model cannot rely on temporal redundancy. The decoder applies an additional **running cell mask** at 50%, further restricting which tokens are reconstructed. This aggressive masking forces the encoder to learn compact, informative representations.

#### Training Procedure

The model was optimized with AdamW [19] ($\beta_1$=0.9, $\beta_2$=0.95) using a base learning rate of $1 \times 10^{-3}$ with 20 epochs of linear warmup followed by cosine decay, for 100 total epochs with a batch size of 40. Input videos were constructed by sampling 16 frames at a stride of 3 from the cine sequences and resized to $128 \times 128$ spatial resolution. The cardiac MRI data is single-channel (grayscale); the channel dimension was replicated to 3 to match the standard architecture. The reconstruction target is mean squared error over masked spatiotemporal patches.

Separate models were trained for the short-axis and long-axis cardiac views using the same architecture and hyperparameters.

##### Embedding Extraction

At inference, all spatiotemporal tokens are passed through the encoder with no masking applied. To account for temporal sampling variability, each video is encoded at multiple temporal offsets and the resulting token sequences are averaged. A single 768-dimensional embedding per scan is obtained by global average pooling across all 2,048 spatiotemporal tokens.

#### S10 ECG Self-Supervised Representation Model (ST-MEM)

For the 12-lead resting ECG data, we employed the Spatiotemporal Masked ECG Modeling (ST-MEM) framework [23], a masked autoencoder adapted for multi-lead electrocardiogram signals.

##### Architecture

ST-MEM extends the masked autoencoder paradigm to handle the multi-lead structure of clinical ECGs. Each of the 12 leads is independently tokenized by a shared 1D patch embedding layer with patch size 75 (corresponding to 0.3 s at 250 Hz sampling rate), producing 30 temporal patches per lead. To encode lead identity, two mechanisms are employed: (1) learnable lead-specific embeddings added to each lead's tokens, and (2) shared learnable separator (`[SEP]`) tokens prepended and appended to each lead's patch sequence. The resulting tokens from all 12 leads are concatenated into a single sequence of $12 \times (30 + 2) = 384$ tokens, which is processed jointly by a standard ViT-B transformer encoder ($d$=768, 12 blocks, 12 attention heads, MLP ratio 4). This design allows cross-lead attention, enabling the model to learn inter-lead relationships. Fixed 1D sinusoidal positional embeddings encode temporal position within each lead.

The decoder is lightweight: embedding dimension 256, 4 transformer blocks, and 4 attention heads. Crucially, the decoder operates *lead-wise*—each lead's tokens (including restored mask tokens) are decoded independently, and reconstruction targets are the raw voltage values within each patch. Per-patch pixel normalization [18] is applied to the reconstruction targets.

##### Training

The model was trained on UK Biobank 12-lead ECG recordings using 75% random masking applied independently per lead. Input signals were randomly cropped to 2250 samples (9 s), bandpass filtered (0.67–40 Hz), and standardized. Training used AdamW [19] ($\beta_1$=0.9, $\beta_2$=0.95, weight decay = 0.01) with a base learning rate of $1.5 \times 10^{-4}$, 40 epochs of linear warmup followed by cosine decay, for 800 total epochs with a batch size of 256.

#### Embedding Extraction

At inference, the full ECG signal is passed through the encoder with no masking. After removing the `[SEP]` tokens, the patch embeddings are averaged across both leads and temporal positions, yielding a single 768-dimensional representation per recording.
